# Sex differences in genetic pathways underlying ischaemic heart disease–depression comorbidity

**DOI:** 10.64898/2026.04.29.26352017

**Authors:** Cato Romero, Alexey A. Shadrin, Nadine Parker, Sara E. Stinson, Linn Rødevand, Dennis van der Meer, Kevin S. O’Connell, Eva van Walree, Jeanne E. Savage, Danielle Posthuma, Ole A. Andreassen, Sophie van der Sluis

## Abstract

**Background and Aims:** Comorbidity between ischaemic heart disease (IHD) and depression (DEP) is prevalent and more pronounced in woman than in men. The biological basis of these sex differences, however, remains unclear. We aimed to assess the contribution of genetic and biological risk factors to sex differences in IHD–DEP comorbidity.

**Methods:** We analysed sex-stratified genome-wide association study summary statistics from 1.14 million individuals of European ancestry across multiple large-scale cohorts and international consortia. Global and local genetic correlations (*r*_g_), pleiotropic loci, and IHD-DEP shared genes were identified using LDSC, MiXeR, LAVA, conjunctional FDR, and FLAMES. We conducted conditional analyses using genetic and phenotypic data for 331 putative risk factors.

**Results:** The *r*_g_ between IHD and DEP was twice as high in females (*r*_g_ =.43) compared to males (*r*_g_ =.21), explaining a greater proportion of comorbidity in females (21% vs 13%). Pleiotropy analyses identified sex-specific genomic regions and genes contributing to IHD–DEP comorbidity. Genetic conditional analysis indicated that behavioural traits (alcohol use, insomnia, social deprivation) contributed more to male IHD–DEP comorbidity, whereas asthma and female-specific health traits contributed more to female IHD–DEP comorbidity. Phenotypic mediation largely reflected the same pattern.

**Conclusions:** Higher IHD-DEP comorbidity in females compared to males is partly attributable to greater shared genetic liability. Distinct genes and differing contributions of behavioural, metabolic, immunological, and reproductive factors further shape these sex differences. These results support sex-aware risk stratification—targeting alcohol, sleep, and loneliness in males and endocrine status and asthma control in females.

---

Women with ischaemic heart disease (IHD) experience higher mortality, worse recovery, and poorer quality of life than men, yet diagnostic criteria, risk prediction, and therapeutic strategies remain largely optimised for male disease presentation and risk profiles (1–4). Individuals diagnosed with depression (DEP) have a higher incidence of IHD, and this association is more pronounced in women (hazard ratios: 1.52 to 3.43) compared to men (hazard ratios: 1.16 to 1.77) (5–10). In addition, DEP is linked to higher rates of hospital readmission and recurrent cardiovascular events, as well as reduced quality of life in IHD patients (11). Notably, DEP contributes to a higher mortality in women with IHD compared to men (12). The higher prevalence and greater clinical severity of IHD–DEP comorbidity in women underscores the need to understand how the mechanisms linking these disorders differ by sex.

Sex differences in IHD-DEP comorbidity suggest that there may also be differences in underlying psychosocial, cardiometabolic, inflammatory, hormonal, and autonomic pathways (13). Because both IHD and DEP show substantial heritability, genetic analyses can provide an exploratory framework to identify the shared biological mechanisms underlying their co-occurrence. Prior genetic studies using sex-combined data have implicated single nucleotide polymorphisms (SNPs) and genes shared between IHD and DEP in pathways related to inflammation, metabolism, psychosocial and lifestyle risk factors (14). However, sex-combined analyses may obscure sex differences in genetic effects that underlie the stronger comorbidity observed in women, highlighting the importance of sex-stratified analyses. To date, only three sex-stratified studies of IHD–DEP have been conducted: a twin study reporting a higher genetic contribution to IHD–DEP comorbidity in female than in male twin pairs (15), and two polygenic score studies showing that genetic risk for DEP predicts IHD events more strongly in women than in men (16,17). These initial results suggest that genetics contributes differently to the IHD–DEP relationship in men and women.

Here, we leverage sex-stratified genome-wide association studies (GWAS) based on 1,144,822 participants (56.4% female) from multiple large-scale cohorts and international consortia to assess sex differences in genetic overlap between IHD and DEP. Through genetic correlations, pleiotropy analyses, gene prioritization analyses, and conditional genetic analyses, we aimed to identify shared and distinct genetic effects and to test whether psychosocial, cardiometabolic, inflammatory, and hormonal risk factors differentially contribute to the IHD–DEP relationship in men and women.

## Methods and Materials

### Participants and phenotype description

Data were analysed from three large population-based biobanks and one international research consortium: UK Biobank (18) (UKB), All of Us Research Program (19) (All of Us), Psychiatric Genomics Consortium (20) (PGC), and Integrative Psychiatric Research (21) (iPSYCH). GWAS data for DEP were available from All of Us, UKB, iPSYCH, and PGC, whereas IHD data were available from All of Us and UKB. Cohort-specific and total sample sizes are shown in **Supplementary Table 1**. Sex was determined based on chromosomal composition, classifying individuals with XX chromosomes as female and those with XY chromosomes as male.

For IHD, UKB cases were identified through ICD-10 codes I21–I25 across primary, hospital inpatient, and death registry. In All of Us, IHD cases were defined by SNOMED CT ID 414545008 (ICD-10 I20–I25) from electronic health records (EHRs).

In All of Us, DEP cases were defined by SNOMED CT ID 370143000 (ICD-10 F32–F33) from EHRs. For UKB, we used sex-stratified summary statistics from Silveira et al. (22), where DEP was defined by ICD-10 (F32, F33, F34, F38, F39) or by self-reported visits to a general practitioner or psychiatrist for depression (fields 2090, 2010). Meta-analysed summary statistics for iPSYCH and PGC were obtained from Blokland et al. (23); iPSYCH diagnoses followed ICD-10 from the Danish Psychiatric Register, and PGC cohorts primarily used DSM-5 diagnoses. All cohorts were restricted to participants of European (EUR) ancestry, based on genetic information. Further cohort details are provided in **Supplementary Note 1**.

### Data acquisition and genome-wide association analysis

We conducted sex-stratified genome-wide association analysis (GWAS) on IHD (UKB, All of Us) and DEP (All of Us), as no public EUR sex-stratified results were available. In All of Us, genotypes were derived from whole-genome sequencing (WGS); SNPs were filtered for MAF <0.01 and missingness >0.05, leaving 7.1M SNPs, and related individuals were excluded (see All of Us pipeline (19)). Association analyses used PLINK(v2.0) (24) with age and the first 16 genetic ancestry PCs as covariates. In UKB, SNP filters were identical with the addition of INFO > 0.9 to account for imputation from SNP arrays, yielding ∼8.5M SNPs. Association analyses used REGENIE(v3.4.1) (25) with Firth penalization and adjustment for genetic relatedness, including age, array, and 20 genetic ancestry PCs as covariates.

### Meta-analysis of DEP and IHD cohorts within males and females

Sex-specific GWAS summary statistics for IHD and DEP were meta-analysed across independent cohorts using sample-size–weighted fixed-effects meta-analysis in METAL(v2020.05.05) (26); IHD data included All of Us and UKB; DEP data included All of Us, UKB, iPSYCH, and PGC. SNPs were aligned to hg19 (GRCh37) using dbSNP (27), and effect alleles were harmonized prior to meta-analysis. Genomic control was not applied because contributing GWAS were already corrected. SNPs with effective sample size <60% of the total were excluded. To retain comparability across traits and sexes, only SNPs present in both sex-stratified IHD and DEP GWAS were kept, yielding 6,040,319 SNPs across 22 autosomes and the X chromosome for downstream analyses.

### Global genetic correlation and contribution to phenotypic relationship

We estimated liability scale SNP-based heritability (*h^2^_SNP_*), genomic inflation (λ_GC_), and sex-stratified genome-wide genetic correlations (*r*_g_) between IHD and DEP using LD score regression (LDSC-v.1.0.1) (28). For liability-scale conversion of *h^2^_SNP_*, sample prevalence was 0.5 due to using effective sample size (as recommended (29)), and population prevalence of IHD was 0.13 and 0.21 for females and males, respectively, and for DEP 0.18 and 0.12 for females and males, respectively. Sex differences in *r*_g_ and *h^2^_SNP_* were tested in Genomic Structural Equation Modelling (gSEM-v.0.0.5) (30) using a model-based Wald test. To assess specificity, we compared IHD with other psychiatric traits and DEP with other cardiovascular diseases using publicly available sex-stratified GWAS (31–35). Analyses used the 1000 Genomes (v3) EUR LD panel and were filtered to HapMap3 SNPs. LDSC provides *r*_g_ estimates corrected for sample overlap.

The genetic contribution to IHD–DEP comorbidity was quantified as the ratio of LDSC-derived genetic covariance to the phenotypic correlation (36,37). Sex-stratified tetrachoric correlations between IHD (ICD-10: I21–I25) and DEP (ICD-10: F32–F34, F38, F39) were estimated in UKB, with standard errors derived using 1,000 nonparametric bootstrap resampling with replacement. Sex differences in phenotypic association were tested using modified Poisson regression with a log link and robust sandwich standard errors (R package *sandwich* v3.1-1), modelling IHD as a binary outcome, DEP as the exposure, and including age as a covariate. A sex-by-DEP interaction term to obtain sex-specific relative risk (RR) and their ratio (RRR) was used. Sensitivity analyses using logistic regression yielded consistent results.

### MiXeR analysis of shared genetic architecture

To estimate the proportion of trait-influencing SNPs that IHD and DEP have in common, we applied bivariate MiXeR (38) v.1.3 to the sex-stratified GWAS data. MiXeR models the genetic architecture of complex traits by assuming a mixture of trait-influencing and non-causal SNPs, following distinct Gaussian distributions. The model accounts for LD structure, genomic inflation due to cryptic relatedness, MAF, GWAS sample size, and sample overlap. The 1000 Genomes (v3) EUR reference panel was used for allele-frequency correction. The MHC region (hg19: chromosome 6, 28,477,797–33,448,354 (39)) was removed to avoid bias from complex LD. Model fit was evaluated using AIC and BIC, with positive values indicating sufficient model fit (40).

### Local genetic correlation analysis across the genome

We estimated local *r*_g_ between IHD and DEP in sex-stratified GWAS using Local Analysis of [co]Variant Annotation (LAVA-v.0.1.0) (41). Unlike LDSC and MiXeR, which estimate genetic overlap across the whole genome, LAVA aims to identify genomic regions where shared genetic signal is strongest. The genome was partitioned into 1,133 ∼2 Mb semi-independent blocks using the LAVA block partitioning algorithm (min. SNPs =5,000), identical settings as in the latest implementation by the authors of LAVA (42), with 1000 Genomes (v3) EUR as LD reference. LAVA accounts for LD within blocks and adjusts for sample overlap using LDSC intercepts.

We first tested univariate local *h^2^_SNP_* within IHD and DEP; only regions with significant genetic signal (LAVA default *P*<1×10□□) in both traits were tested for bivariate *r*_g_. This threshold filters out regions with low genetic signal while retaining a substantial proportion of sub-genome-wide significant loci that may contribute to the global *r*_g_. Remaining regions were then tested for bivariate local *r*_g_, applying multiple testing correction (α*_BON□_* =.05/(74+95) =2.96×10^−4^; two-sided).

To test for statistical independence of significant male and female local *r*_g_, we performed partial local *r*_g_ analysis, conditioning IHD–DEP correlations in one sex on the corresponding genetic signal in IHD and DEP of the other sex within the same locus. Significant partial *r*_g_ indicated genetic overlap unique to that sex (α*_BON□_*=.05/number of LAVA loci tested in sensitivity analysis =.05/8 =6.25×10^−3^; two-sided). Loci lacking univariate signal for both traits in one sex were classified as sex-specific for the opposite sex.

### Pleiotropic SNP detection using conjunctional false discovery rate

Pleiotropic loci shared between IHD and DEP were identified using the conjunctional FDR (conjFDR) framework (43). Conditional FDR (cFDR) quantifies the probability a SNP is null for one trait given its association with the other, based on enrichment of low P-values across quantile bins. A lower cFDR value indicates a lower probability of the SNP being a false positive and therefore higher confidence that it represents a true association. ConjFDR was defined as the maximum of cFDR_DEP|IHD_ and cFDR_IHD|DEP_, and SNPs with conjFDR < 0.05 were considered jointly associated. Loci were defined by the span of significant SNPs. Sex-stratified GWAS and the 1000 Genomes (v3) EUR reference were used as input.

### Gene prioritization of shared IHD–DEP loci

Gene prioritisation was performed within pleiotropic loci identified by either LAVA or conjFDR. Several LAVA and conjFDR loci were relatively large for gene prioritization (>2Mb). To circumvent this we split each locus into LD-independent subsets. Specifically, within each male and female locus, SNPs were first clumped into ∼LD-independent subsets using PLINK(v.1.9; --clump-kb 250, --clump-p1 1; --clump-r2 0.1), done separately in IHD and DEP and merged when regions overlapped. To prioritize likely causal, or *effector*, genes within pleiotropic regions, we applied Fine-mapped Locus Assessment Model of Effector geneS (FLAMES-v1.1.0). FLAMES is a machine-learning-based prediction model that integrates SNP fine-mapping results with functional genomic annotations to identify genes most likely mediating trait associations. FLAMES requires two inputs: (i) credible SNP sets likely to contain the causal variant within each region, and (ii) gene-level polygenic prioritisation scores (PoPS-v.0.2; (44)). To derive the credible SNP sets, we fine-mapped each pleiotropic LAVA/conjFDR locus for IHD and DEP separately using SuSiE (v.0.12.41) (27), assuming one causal SNP per locus since generating multiple credible sets requires individual-level data that were unavailable for all cohorts. For each locus and trait, we then obtained a 95% credible set, defined as the smallest set of SNPs with a 95% probability of containing the causal variant. We then ran PoPS with co-expression features using output from MAGMA (45) gene-based test and gene property tissue analysis using GTEx(v.8). FLAMES estimated the cumulative precision value of each gene within the locus, applying the default precision threshold of 0.75. Genes were considered shared effector genes if they were predicted in both IHD and DEP within the same locus with a score above 0.75. LocusZoom plots were generated for shared gene loci using LDlink (46).

### Conditional genetic correlation analysis of shared IHD–DEP risk factor traits in Genomic SEM

To test whether sex-specific global *r*_g_ between IHD and DEP was mediated by shared risk factor traits we conducted conditional *r*_g_ analysis in gSEM. We evaluated 331 potential risk factor traits using sex-stratified GWAS data, selected based on biological relevance to cardiovascular, metabolic, inflammatory, hormonal, or behavioural pathways, as well as prior epidemiological or genetic evidence linking them to IHD or DEP (47–49). The risk factor set included anthropometric traits (e.g., BMI), social measures (e.g., Townsend deprivation index), 249 metabolic (50) and inflammatory markers (e.g., cholesterol, C-reactive protein), and lifestyle or clinical factors (e.g., smoking, alcohol use behaviours, insomnia).

Implemented in gSEM, the conditional *r*_g_ analysis compares the marginal *r*_g_ between two traits (Y, X_1_) with the conditional *r*_g_ estimate after adjusting for a risk factor trait (X_2_). A significant difference between the marginal and conditional *r*_g_ (marginal *r*_g_ ≠ conditional *r*_g_) indicates that X_2_ contributes significantly to the genetic covariance between Y and X_1_. From the conditional analyses we derived the following estimates: *direct effect*, which is the residual *r*_g_ between X_1_ and Y after conditioning of X_2_; *indirect effect*, which is the product of the *r*_g_ between the predictors (X_1_, X_2_) and the residual *r*_g_ between X_2_ and Y after conditioning of X_1_; *total effect*, which is the sum of the direct and indirect effect; and *proportion mediated*, which is the proportion of *indirect effect* to the *total effect.* Risk factor traits were tested only if they showed a nominally significant *r*_g_ with both IHD and DEP in either sex (*P* < 0.01). Significant conditional effect of risk factor traits was determined by *P*-value of the residual *r*_g_ between X_2_ and Y after conditioning of X_1_ (α_BON_ =.05/number of traits tested =.05/204=2.45×10^−4^); see (51) for details). To avoid bias from directional model specification, conditional effects were estimated with both possible outcomes (IHD→DEP and DEP→IHD). Conditional effect often appears inflated when the outcome is the trait most strongly correlated with the risk factor trait. By comparing both model orientations and reporting the one with the smaller indirect effect, we ensured a conservative and robust estimate of mediated genetic overlap.

### Conditional phenotypic analysis in UKB

To test whether conditional genetic associations aligned with phenotypic patterns, we conducted sex-stratified conditional analyses in the UK Biobank. Analyses were run separately for each risk factor trait that showed significant conditional genetic effects. We implemented counterfactual mediation models using the *regmedint* R package (52) to derive direct effect, indirect effect, total effect estimates, as well as proportion mediated for each risk factor trait. Similarly to the conditional genetic analysis, phenotypic analyses were run in both directions of association (i.e., IHD→DEP and DEP→IHD), and we report the pathway with the lowest proportion mediated as a conservative estimate. Logistic regression was used and always included age as a covariate. When mediation by blood derived measurements were assessed, statin use (field 20003; atorvastatin, rosuvastatin, simvastatin) was included as additional covariate. Significant conditional effects of risk factor traits were corrected for multiple testing (α_BON_ =.05/number of traits tested =.05/34 =1.47×10^−3^).

## Results

### Sex differences in genetic and phenotypic overlap between IHD and DEP

We conducted a GWAS meta-analysis of sex-stratified summary statistics of IHD and DEP using METAL, followed by heritability assessment using univariate LDSC (**Methods;** for cohort sample sizes see **Supplementary Table 1**; for cohort descriptions see **Supplementary Note 1**). Effective sample sizes differed by sex (DEP_male_ =174,413 vs. DEP_female_ =282,329; IHD_male_ =149,866 and IHD_female_ =87,986), reflecting ≈62% larger DEP samples in females and ≈70% larger IHD samples males. All GWAS showed appropriate inflation metrics, reflecting polygenic architecture rather than bias (**Supplementary Table 2**). Heritability from common SNPs (*h*^2^_SNP_) was higher in IHD_male_ (*h*^2^_SNP_ =.15, SE =.01) than in IHD_female_ (*h*^2^_SNP_ =.12, SE =.01; difference =.03, SE =.01, *P* =.004), reflecting a modest yet statistically significant difference, whereas DEP_male_ (*h*^2^_SNP_ =.07, SE =.004) and DEP (*h*^2^_SNP_ =.07, SE =.003; difference =.00, SE =.01, *P* =.54) were comparable.

Global genetic correlation (*r*_g_) between IHD and DEP was substantially higher in females (IHD_female_–DEP_female_: *r*_g_ =.43, SE =.04, *P* =4.06×10^−28^) than in males (IHD_male_–DEP_male_: *r*_g_ =.21, SE =.03, *P* =3.22×10^−9^; difference =.22, SE =.05, *P* =1.94×10□^5^; **Supplementary Table 3**; **Methods**), while cross-sex *r*_g_ estimates (i.e., IHD_male_–DEP_female_ vs. IHD_female_–DEP_male_) did not differ, suggesting that the sex difference in IHD–DEP *r*_g_ cannot be attributed to one trait alone. Sensitivity analyses confirmed that the sex difference in IHD–DEP *r*_g_ was robust to potential confounds including shared-sample overlap, sample size imbalance, and heterogeneity in IHD and DEP phenotype definitions (**Supplementary Note 2**).

To assess whether the sex difference in the global *r*_g_ between IHD and DEP reflects a disorder-specific or broader pattern, we extended the sex-stratified global *r*_g_ analyses to 7 additional psychiatric and 7 additional cardiovascular disorders (see **Supplementary Table 4** for overview). For IHD, a sex difference was observed only for ADHD, with higher *r_g_* with IHD for female than male individuals, but not for other psychiatric phenotypes (**Figure 1A**). For DEP, nominally higher *r_g_*s in females were seen with heart failure and conduction disorder, but not with the other cardiovascular diseases, and these did not remain significant after multiple testing correction (**Figure 1B**). Thus, apart from ADHD, sex differences in global *r*_g_ appears largely confined to the IHD–DEP relationship.

**Figure 1.**
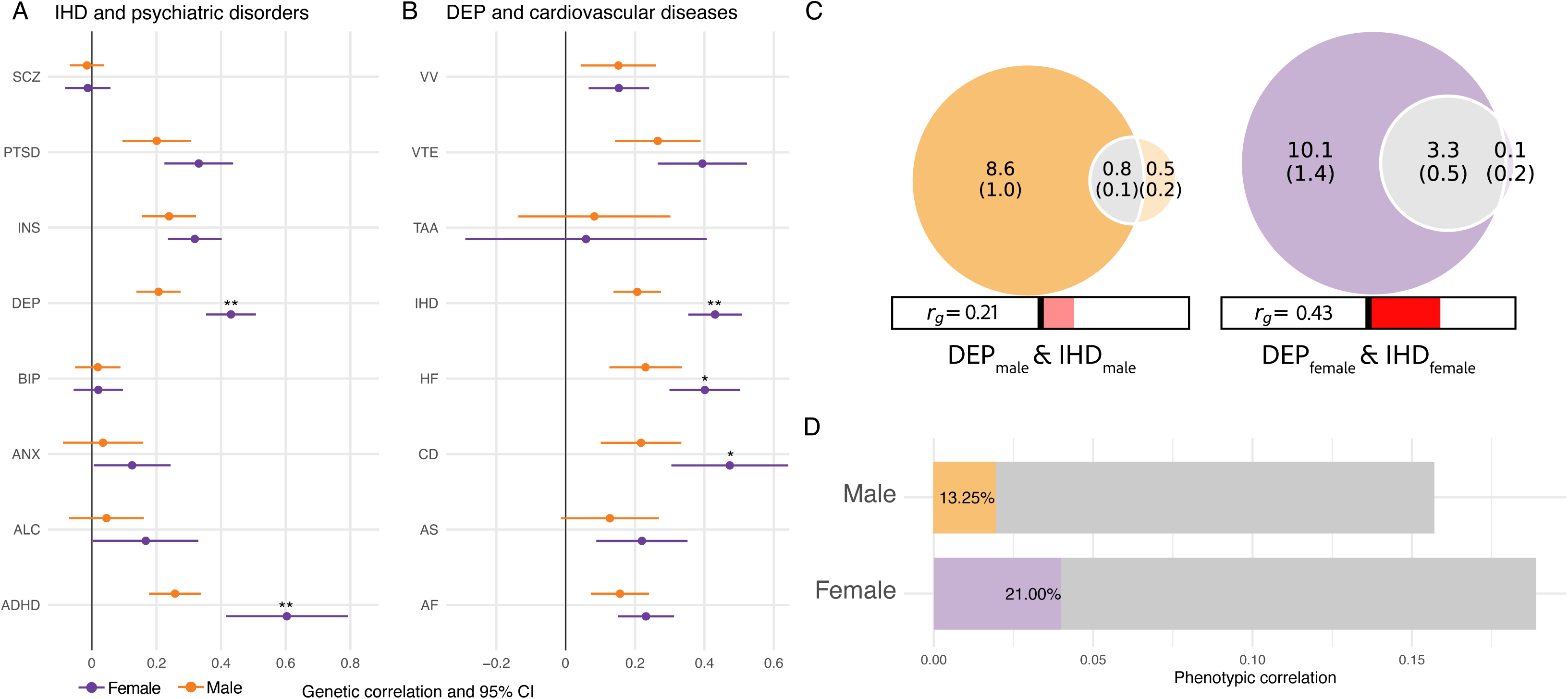
Genetic overlap between IHD–DEP is larger in females than in males. A) LDSC global r_g_ between IHD and eight psychiatric disorders in males (orange) and females (purple). SCZ =schizophrenia; PTSD =post-traumatic stress disorder; INS =insomnia; BIP =bipolar disorder; ANX =anxiety disorder; ALC =alcohol dependency; ADHD =attention-deficit hyperactivity disorder. B) Global r_g_ between DEP and eight cardiovascular diseases in males and females. AF =atrial fibrillation; AS =aortic valve stenosis; CD =conduction disorder; HF =heart failure; TAA =thoracoabdominal aneurysm; VTE =venous thromboembolism; VV =varicose veins. Correlation estimates are plotted with 95% confidence intervals. ‘*’ indicates a sex difference in r_g_ for the same trait at P < 0.05, ‘**’ at Bonferroni adjusted P < 0.05. C) Venn diagram of estimated bivariate MiXeR shared genetic architecture between IHD (right circle) and DEP (left circle) in males (orange) and in females (purple). Estimates are in 1000 SNPs, i.e., 3.3 (.5) denotes 3,300 shared SNPs with SE =500 between IHD and DEP in females. Bar under Venn diagram displays the r_g_ estimate. MiXeR model fit was supported by AIC, but not BIC in both sex-stratified analyses, indicating pleiotropic overlap whose precise magnitude will need confirmation in higher-powered GWAS. D) Tetrachoric correlations between IHD and DEP in males and females (grey bar) with the coloured bars (male =orange, female =purple) showing the proportion of comorbidity that can be attributed to genetic covariance.

To corroborate the sex difference in global *r*_g_ between IHD and DEP, we applied MiXeR (**Methods**), which quantifies shared genetic architecture as the proportion of estimated trait-influencing SNPs in common. Bivariate MiXeR estimated ∼800 (SD =100) trait-influencing SNPs shared between IHD_male_ and DEP_male_ (Dice coefficient =.18), and ∼3,300 (SD =500) trait-influencing SNPs shared between IHD_female_ and DEP_female_ (Dice coefficient =.65; **Figure 1C; Supplementary Table 5**), suggesting that the genetic overlap between IHD and DEP is larger in females than in males, both in magnitude and in the number of trait-influencing SNPs shared.

We next examined to what extent the observed genetic overlap contributes to sex differences in phenotypic comorbidity between IHD and DEP. In the UK Biobank, the phenotypic correlation (*r*) was .19 among women and .16 among men (*r*-difference =.03, SE =.01, *P* =2.21×10□□), corresponding to a 24% higher relative risk of IHD among individuals with DEP in women (RR =1.97, 95% CI =[1.90, 2.03]) than in men (RR =1.58, 95% CI =[1.54, 1.62]; RRR =1.24, 95% CI =[1.19, 1.29]; **Methods**). The proportion of genetic covariance (LDSC) relative to the phenotypic correlation—reflecting the fraction of phenotypic comorbidity explained by shared genetic liability—was estimated at 21% for women and 13% for men (**Figure 1D**). In summary, the larger genetic overlap between IHD and DEP in women accounts for a greater fraction of their phenotypic comorbidity than in men.

### Sex-specific genetic effects underlying comorbidity between IHD–DEP

To identify SNPs with sex-specific effects (that is, SNPs contributing to comorbidity in only one sex) or sex-dependent effects (that is, SNPs contributing to comorbidity in both sexes but with different effect sizes), we applied several approaches. Firstly, we estimated local genetic correlations (local *r*_g_s) using LAVA (**Methods**) to identify genomic regions harbouring shared genetic signal between IHD and DEP in males and females separately. After filtering on univariate genetic signal, 74 and 95 of the 1,133 predefined, roughly independent genomic regions remained for males and females, respectively, with 11 regions overlapping between sexes (more regions overlapping than expected by chance; Fisher’s exact test OR =2.03, *P* =.04). Bivariate local *r*_g_ identified one significant region for IHD–DEP in males and seven significant regions in females after multiple testing correction (**Supplementary Table 6–7; Supplementary Figure 1A–B**). All significant local *r*_g_s were positive, suggesting concordant, risk-increasing effects for IHD and DEP, and no locus harboured significant local *r_g_* for both sexes. Sensitivity analysis indicated that most associations (6/8) were sex-specific rather than sex-dependent (**Table 1**).

**Table 1.**
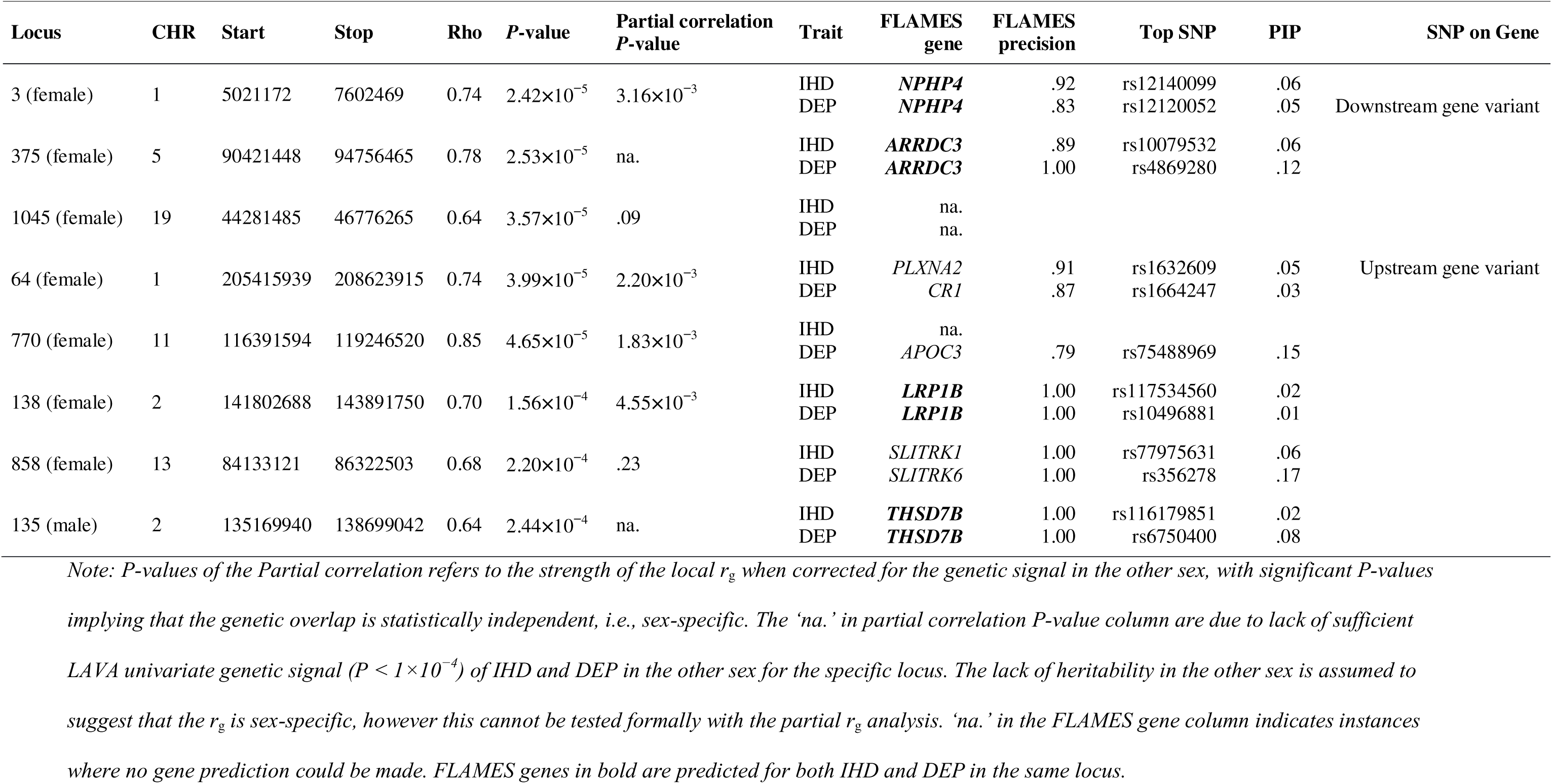
Genomic regions showing significant local genetic correlation between IHD and DEP in males and females.

Secondly, to identify specific loci jointly associated with IHD and DEP, we applied the conjFDR approach (**Methods**). In males, we detected five independent loci shared between IHD and DEP, and in females we detected four independent loci shared between IHD and DEP (**Table 2; Supplementary Table 8–9; Supplementary Figure 2A–B**). In all loci, the direction of SNP effects was the same for IHD and DEP, and no loci overlapped between males and females.

**Table 2.** Genomic regions showing <.05 conjFDR pleiotropic association between IHD and DEP.

| Sex | CHR | Start | Stop | conjFDR<br>P-value | Opposite sex<br>conjFDR P-value | Trait | FLAMES<br>gene | FLAMES<br>precision | Top SNP | PIP | SNP on Gene |
| --- | --- | --- | --- | --- | --- | --- | --- | --- | --- | --- | --- |
| Female | 3 | 48964303 | 50067450 | .003 | .25 | IHD<br>DEP | na.<br>na. |  |  |  |  |
| Female | 4 | 2872507 | 3499521 | .022 | .92 | IHD<br>DEP | **HTT**<br>**HTT** | .80<br>.78 | rs4690072<br>rs3095075 | .13<br>.10 | Intron variant<br>Downstream gene variant |
| Female | 8 | 19679778 | 20179778 | .046 | .45 | IHD<br>DEP | *LPL*<br>*ATP6V1B2* | 1.00<br>0.79 | rs268<br>rs9644568 | .67<br>.05 | Missense variant<br>Upstream gene variant |
| Female | 14 | 75110906 | 75610815 | .021 | .30 | IHD<br>DEP | na.<br>na. |  |  |  |  |
| Male | 3 | 48481487 | 50450789 | .009 | .53 | IHD<br>DEP | na.<br>na. |  |  |  |  |
| Male | 7 | 12020815 | 12521136 | .009 | .93 | IHD<br>DEP | *TMEM106B*<br>*ARL4A* |  | rs6460894<br>rs10234805 | .04<br>.03 | Upstream gene variant<br>Upstream gene variant |
| Male | 7 | 113841844 | 117807051 | .013 | .51 | IHD<br>DEP | **FOXP2**<br>**FOXP2** | 1.00<br>.98 | rs10262103<br>rs6942634 | .13<br>.26 | Intron variant<br>Upstream gene variant |
| Male | 12 | 56733425 | 57581271 | .048 | .94 | IHD<br>DEP | na.<br>na. |  |  |  |  |
| Male | 12 | 121166988 | 122208610 | .031 | .98 | IHD<br>DEP | *HNF1A*<br>na. |  | rs2244608 | .46 | Intron variant |

Note: 'na.' in the FLAMES gene column indicates instances where no gene prediction could be made. FLAMES genes in bold are predicted for the both IHD
and DEP in the same locus. 'Opposite sex conjFDR P-value' refers to the conjFDR p-value of the same loci in the other sex.

We next used FLAMES (**Methods**) for gene prioritization to identify candidate effector genes underlying the specific pleiotropic genomic regions detected with LAVA or conjFDR (MAGMA input files are provided in **Supplementary Tables 10–11**). For males, two pleiotropic genes were identified—*THSD7B* (from a LAVA locus) and *FOXP2* (from a conjFDR locus). For females, four pleiotropic genes were identified—*LRP1B, NPHP4, and ARRDC3* (from LAVA loci), and *HTT* (from a conjFDR locus; **Table 1–2; Supplementary Table 12;** for locus zoom plots see **Supplementary Figure 3–8**). The identified genes represent plausible biological mediators of sex-specific comorbidity patterns.

### Assessment of sex-shared and sex-specific risk factor traits using conditional genetic analysis

Sex differences in shared genetic liability, comorbidity, and locus-specific effects allow for the possibility that risk factors contribute differentially between sexes to the IHD–DEP relationship. To examine this, we applied conditional *r*_g_ analysis in Genomic SEM (**Methods**). Of the 331 candidate risk factors (full list in **Supplementary Table 13**), 204 were genetically correlated with both IHD and DEP in either sex and subsequently included in the conditional analysis (see **Supplementary Note 3** for a detailed example).

Genetic conditioning on personal history of alcoholism produced the strongest reductions in *r*_g_ between IHD and DEP, reducing to non-significant *r*_g_ in males and reducing the *r*_g_ by nearly half in females (**Figure 2)**. Smoking showed consistent though relatively smaller effects across sexes (14% in females, 30% in males). Male-specific influences were observed for loneliness (63%), insomnia (40%), 20 different fatty-acid blood measurements (range 17%–34%), Townsend deprivation index (31%), glycoprotein acetyls blood measurement (28%), and waist-hip ratio (23%), while these traits were not significant for females. Female-specific effects were seen for excessive or irregular menstruation (25%), (age at initiation of) hormone replacement therapy (HRT) use (21%-24%), and asthma diagnosis (23%) (**Figure 2**; for full results see **Supplementary Table 14** and **Supplementary Figure 9**).

**Figure 2.**
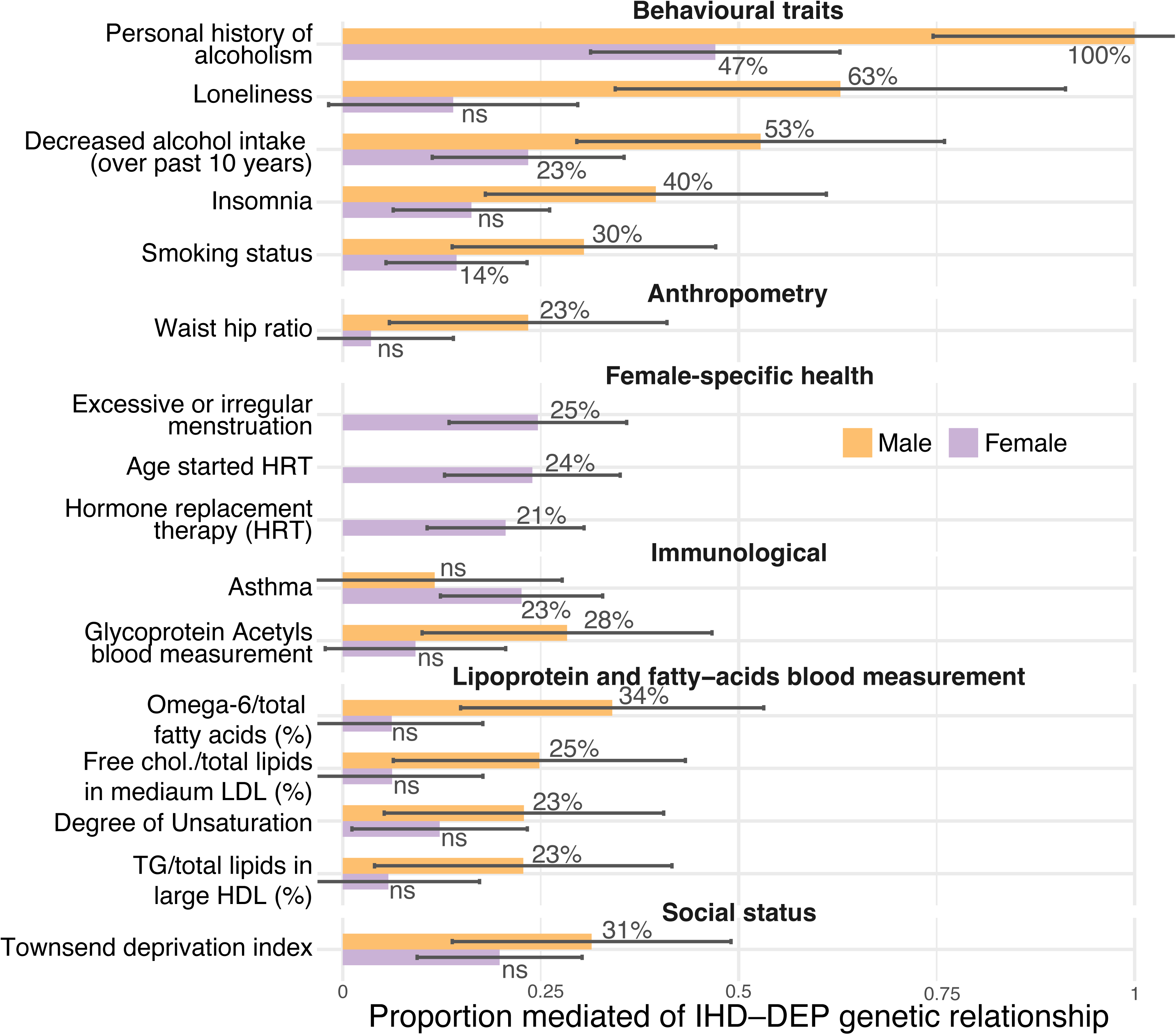
Conditional genetic effect of risk factor traits on IHD–DEP genetic association. Significant genetic conditional traits in either sex are plotted for males (orange) and females (purple). The y-axis shows the proportion mediated, defined as the ratio of the indirect effect of the risk factor trait to the marginal IHD-DEP r_g_. For each risk factor trait, conditional effect was estimated in both model orientations (IHD→DEP and DEP→IHD), and the minimum proportion mediated is plotted as a conservative estimate; maximum proportions (average difference between minimum and maximum =0.062 among significant risk factor traits) are reported in **Supplementary Table 14**. Standard errors of the indirect effects were approximated using the standard errors of the corresponding direct effects, as recommended (79). Female-specific health traits are shown only for females. NS indicates non-significant conditional effect. For lipoprotein and fatty-acid blood measurements, only a representative subset of 20 significant markers is displayed; the full set is shown in **Supplementary Figure 9**.

At the phenotypic level, conditional findings broadly mirrored the genetic results (**Figure 3**). Personal history of alcoholism again showed the strongest mediating effect, significantly more pronounced in men (29.7%) than in women (20.2%). Significantly stronger conditional effects for men were additionally seen for insomnia (9.4% in men, 7.0% in women) and Townsend deprivation index (11% in men, 9% in women), while loneliness (∼10%), smoking (∼7%) and waist-hip ratio (∼15%) had comparable conditional effects in both sexes. Asthma showed a twofold stronger effect in women (8.8%) compared to men (4.2%), and female-specific health traits demonstrated moderate conditional effects, including HRT use (3.9%), age at starting HRT (2.5%), and irregular menstruation (1.4%). Contrary to conditional *r*_g_ results, fatty-acid and glycoprotein acetyl blood measurement conditional results were stronger in women (range 5.7%–10.8%) than in men (range 2.7%–6.8%) (for full results see **Supplementary Table 15**).

**Figure 3.**
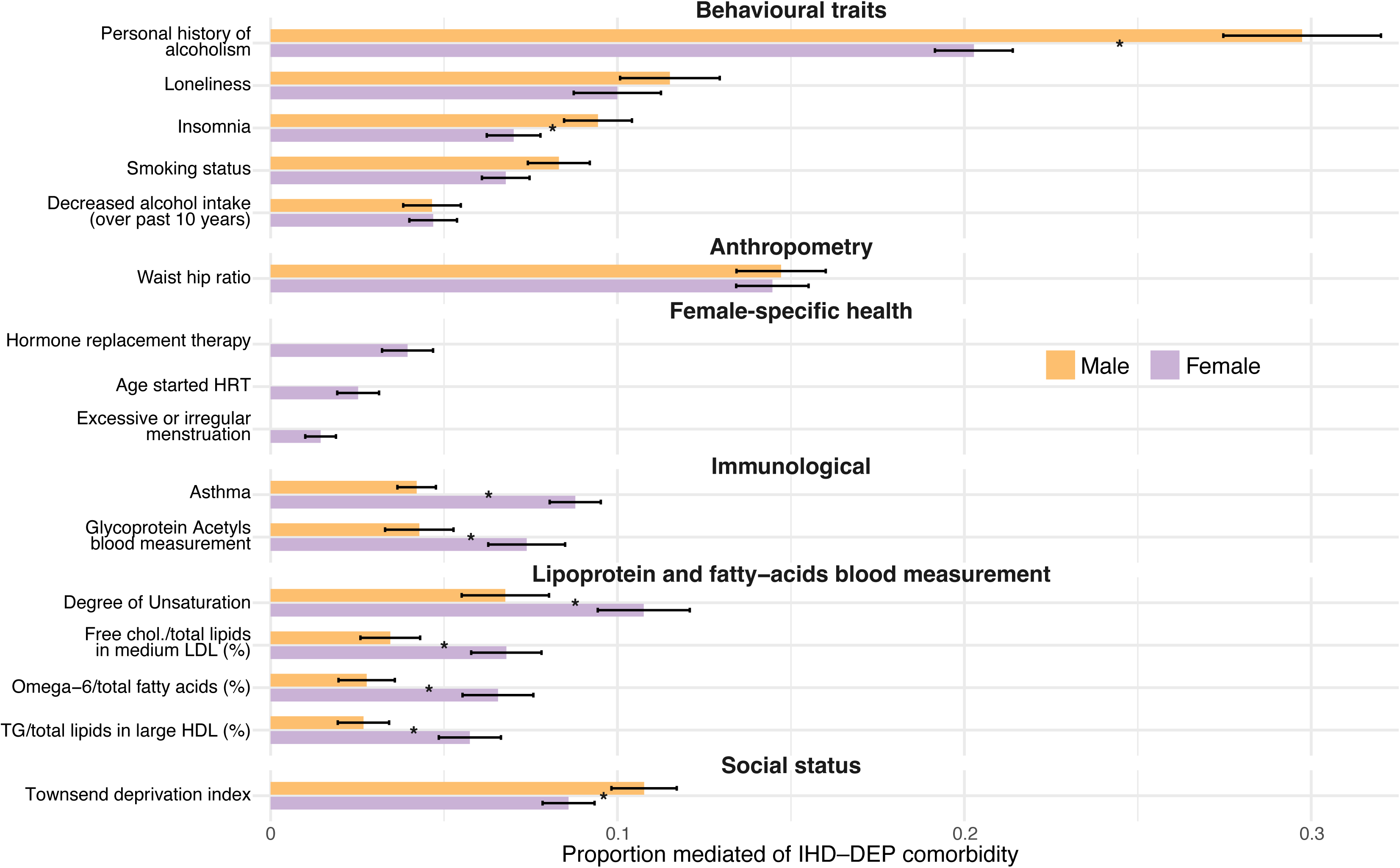
Conditional phenotypic effect of risk factor traits on the IHD–DEP association. Phenotypic conditional effects of risk factor traits are shown for males (orange) and females (purple). The y-axis displays the proportion mediated, defined as the ratio of the indirect effect of the risk factor trait to the marginal phenotypic association between IHD and DEP. For each risk factor trait, conditioning was estimated in both model orientations (IHD→DEP and DEP→IHD), and the minimum proportion mediated is plotted as a conservative estimate; maximum proportions (average difference =0.00318 among significant risk factor traits) are reported in **Supplementary Table 15**. Standard errors of the indirect effects were approximated using the standard errors of the corresponding direct effects, as recommended (79). Differences in proportion mediated between sexes were tested using Wald tests; after correction (αBON =0.05/34), decreased drinking, waist–hip ratio, loneliness, and smoking showed no significant sex differences, while all other risk factor traits did (indicated by * in the plot).

## Discussion

In this study, we observed marked sex differential genetic effects underlying the comorbidity between IHD and DEP, with corroborating phenotypic associations. We found a stronger shared genetic liability in females, with the IHD–DEP genetic correlation approximately twice as high in females (*r*_g_ =.43) as in males (*r*_g_ =.21), and a larger fraction of the phenotypic comorbidity attributable to genetic covariance in females (21% versus 13%). MiXeR similarly estimated more SNPs overlapping between IHD and DEP in females (Dice coefficient =.65) than in males (Dice coefficient =.18). While earlier work has suggested that the IHD–DEP genetic relationship can be sex-dependent at a global level (15–17), our study adds specific genomic loci, identifying candidate effector genes and describing genetic overlap shared with risk factors that may underlie this difference. These findings underscore the importance of sex-stratified genetic analyses for disentangling pathophysiological differences and risk factors for IHD–DEP comorbidity in men and women, and suggest that men and women with comorbid IHD and DEP may benefit from sex-informed clinical monitoring and prevention or treatment strategies.

We identified specific loci and genes that may underlie the sex difference in shared IHD–DEP genetic liability. The set of genes comprised four female-prioritised genes (*LRP1B, HTT, ARRDC3, NPHP4*) and two male-prioritised genes (*FOXP2, THSD7B*; **Table 3**). Data from gene knock-out mouse models were available for four of these genes and three showed sex-specific phenotypes—two female-specific (*LRP1B*(53)*, HTT*(53)) and one male-specific (*THSD7B*(53)), consistent with our human genetic results (**Table 3**). Three of the female-prioritised genes show direct oestrogen-linked regulation (*LRP1B* via oestrogen-linked chromatin accessibility (54,55), *HTT* via oestradiol (56), *ARRDC3* via oestrogen in liver (57)), and both male-prioritised genes relate to androgen-linked regulation (*FOXP2*(58)*, THSD7B*(59)). Reproductive and pregnancy-related biology is prominent in three of four female-prioritised genes (*LRP1B* (60)*, ARRDC3* (61,62)*, NPHP4* (63–66)), including links to menarche and preeclampsia. Cardiometabolic pathways (lipids, BMI, atherosclerosis/IHD risk, autonomic/cardiac conduction) and neurobehavioral/affective associations (neuroticism, anxiety, DEP, ADHD, insomnia, risk-taking) are shared across all six genes. An oestrogen-responsive neuroprotective programming role recurs in both *LRP1B* (APP/Aβ (54,55)) and *HTT* (neuroglobin trafficking (56)), with potential attenuation around menopause.

**Table 3.** Summary of prioritised gene descriptions for females and males.

| Sex | Gene | Expression / Regulation | Sex-specific findings | Functional links | GWAS/traits implicated |
| --- | --- | --- | --- | --- | --- |
| **Female** | *LRP1B* | Brain; oestrogen-regulated chromatin; spliced in adrenal/testis | Female ↑ circulating iron in KO mice | Neuroprotection (↓ amyloid toxicity), food intake | Vaginal microbiome, breastfeeding, menarche, neuroticism, alcohol, preeclampsia, CVD |
|  | *HTT* | Brain; estradiol-regulated neuroprotection (neuroglobin) | Female phenotypes: hyperactivity, grooming, ↓ weight; female-specific CAG cortical effects | Resilience to oxidative stress; cardiac disorders, lipid profiles | BMI, alcohol, worry, DEP |
|  | *ARRDC3* | Liver; oestrogen-regulated | Female offspring growth (methylation); pregnancy-related roles | Insulin/glucose metabolism, β-adrenergic signaling, obesity/energy, angiogenesis, preeclampsia | Obesity, metabolism, mood vulnerability |
|  | *NPHP4* | Ciliary transition-zone; Wnt/Shh signaling | Ovarian dysfunction, placenta retention/abruption | Follicle development, luteinization, steroidogenesis; heart malformations, IHD risk | Cardiovascular events, neuroticism |
| **Male** | *FOXP2* | Brain; androgen-responsive; neural/cardiopulmonary development | KO mice: ↑ respiratory rate, abnormal eating, CV dysfunction, ↑ dopamine/serotonin | Synapse regulation; sinoatrial node development; cortical-autonomic link | DEP, schizophrenia, apraxia, ADHD, insomnia, risk-taking |
|  | *THSD7B* | Cytoskeleton organization; angiogenesis; androgen-regulated | Male phenotypes: ↓ body length, ↑ triglycerides | Angiogenesis, prostate cancer, metabolic traits, ethanol response | Smoking initiation, neuroticism, BMI, lipids |

*Note: ADHD =attention-deficit/hyperactivity disorder; BMI =body mass index; CAG =cytosine-adenine-guanine trinucleotide repeat; CVD =cardiovascular* *disease; KO =knockout; Shh =Sonic hedgehog; Wnt =Wingless/Integrated signalling pathway; ↑ =increased; ↓ =decreased. Detailed description, discussion* *and sources in **Supplementary Note 4.***

Overall, the female-prioritised genes converge on oestrogen-responsive neuroprotection and reproductive/pregnancy pathways with cardiometabolic overlap, whereas the male-prioritised genes converge on androgen-linked neurodevelopmental and metabolic–behavioral liability with autonomic/cardiac features (for a full discussion, see **Supplementary Note 4**).

In our conditional *r*_g_ analyses, personal history of alcoholism had the largest impact: conditioning on this trait reduced IHD–DEP genetic overlap in both sexes—nullifying it in males and markedly attenuating it in females. Decreased alcohol intake over the past 10 years also showed a substantial conditional effect, whereas conditioning on a current alcohol use disorder diagnosis showed no significant effect. This suggests that the strongest effects reflect past significant alcohol problems and potentially medically relevant alcohol pathology (e.g., other medication/treatment interactions) rather than current drinking or alcohol dependency per se. Mechanistically, this aligns with the male-prioritised gene *THSD7B*, which has been linked to heightened ethanol sensitivity, and its knockout induces hypertriglyceridemia in male—but not female—mice, suggesting a cardiometabolic route through which adverse effects of alcohol may be amplified in males. Other traits—loneliness, insomnia, Townsend deprivation index, and waist-hip ratio—reduced the IHD–DEP genetic overlap only in males, pointing to social and behavioural pathways that are more relevant to shared IHD–DEP liability in males. This pattern is consistent with epidemiological observations that environmental risk-factor burdens contribute more strongly to IHD in men than in women (67).

In contrast, asthma showed a larger impact in females in genetic and phenotypic conditional analyses, matching prior reports that IHD risk from adult asthma-onset is only present in women (68), as well as direct links between asthma susceptibility and female-specific hormonal pathways (69,70). A putative link with asthma may involve female-prioritised gene *HTT*, as the top *HTT*-linked SNPs prioritised in the female IHD–DEP analysis have also been associated with asthma (**Table 2**) (71). Female-specific risk factor traits—(age at initiation of) HRT use, irregular or excessive menstruation—further highlight the role of hormonal regulation in female IHD–DEP comorbidity, consistent with evidence that reduction in oestrogen after menopause increased risk for both IHD and DEP (72,73). Genetic conditioning due to HRT use likely reflects the symptoms and hormonal changes that prompt HRT initiation, rather than medication side-effects, suggesting that HRT use primarily indexes underlying endocrine vulnerability. Female-prioritised genes map to oestrogen-responsive neuroprotective programmes (*HTT*, *LRP1B*) and reproductive control (*NPHP4*), providing a putative mechanistic link between female reproductive biology and increased IHD–DEP vulnerability.

Lastly, blood-based measurements of fatty-acids and glycoprotein acetyls showed a complex pattern of sex differences. All blood measures showed genetic conditioning only in males, while phenotypic conditioning indicated a comparatively larger role for the same metabolite and inflammatory markers in females. This may reflect stronger environmental influences on the female serum metabolome—arising from known factors such as diet, physical activity, obesity, sleep, socioeconomic status, hormonal changes, contraceptive use, and education (74). Another explanation is that the genetic liability of female depression includes a stronger metabolic component (22,75), which may distribute genetic effects across a broader metabolic profile, preventing any single metabolite from reaching significance in the conditional *r*_g_ analysis.

Taken together, these observations highlight specific sex differences in the genetic liability shared between IHD and DEP that point to differential involvement of endocrine, inflammatory, cardiometabolic, and behavioural pathways. These results have several potential clinical implications. Current cardiovascular risk prediction algorithms perform less well in women than in men and remain largely calibrated to male-typical risk profiles (76). Our findings suggest that incorporating factors relevant to women—including reproductive history, menopausal status, HRT-related symptomatology, asthma, metabolomic- and inflammatory markers—could strengthen cardiovascular risk models for women and help identify those most likely to develop IHD–DEP comorbidity. For men, behavioural and social factors, such as history of alcohol abuse, smoking behaviour, sleep disturbance, loneliness, and socioeconomic deprivation may further improve risk prediction estimates. In line with recent evidence, several of our sex-specific conditional traits overlap with predictors highlighted in contemporary IHD risk-prediction research—such as triglycerides for women and lipoprotein markers and social deprivation index for men (76). Our findings highlight additional sex-specific factors that may further refine clinical risk tools for identifying individuals vulnerable to IHD–DEP comorbidity.

The study has some relevant limitations. All data were from individuals of European ancestry, limiting generalizability. Phenotypic analyses may also be affected by UK Biobank’s healthy volunteer bias (77), which can influence comorbidity and conditional estimates. The phenotypic definitions of IHD and DEP were broad, and sex differences in IHD comorbidity patterns or unmeasured variables could contribute to differential associations with DEP. Likewise, variation in IHD subtypes (not identifiable through ICD-10 codes alone) may mask subtype-specific relationships with DEP that differ between females and males. Separating sex-specific, sex-dependent, and shared genetic effects relies inherently on statistical power; therefore, the extent to which genetic effects are truly sex-specific must be confirmed in GWAS with larger sample sizes. Moreover, the relatively low *h*^2^_SNP_ (especially of DEP) limits power to detect specific genetic overlap. Given that conjFDR is affected by sample overlap, which is present here, the pleiotropic signals at *HTT* and *FOXP2* require independent replication. Still, locus-zoom patterns support that these associations are likely genuine in both traits. Finally, what appears as sex-specific pleiotropy may in part reflect gene-by-environment interactions due to sex differences across hormonal stages, medications, and healthcare utilization—or ascertainment differences within cohorts (77,78).

Together, these considerations highlight the need for replication in more diverse samples, deeper phenotyping, and longitudinal designs to further clarify the mechanisms underlying sex-differentiated IHD–DEP comorbidity.

## Conclusion

In summary, we find that the genetic overlap between IHD and DEP is substantially larger in females than in males, which is supported by phenotypic associations and is reflected in pleiotropic loci and gene discovery with potentially sex-specific effects. Female-prioritised genes converge on oestrogen-responsive neuroprotection, reproductive biology, and cardiometabolic pathways, whereas male-prioritised genes point toward androgen-linked neurodevelopmental, behavioural, and metabolic liability. Conditional analyses further show that the contributors to shared IHD–DEP risk differ by sex, reflecting a greater role for behavioural and social factors in males and for endocrine and inflammatory pathways in females. These findings motivate sex-aware approaches to risk stratification and prevention of IHD–DEP comorbidity. For men, this may involve attention to behavioural and metabolic profiles; for women, this may involve monitoring hormonal status, menopausal transition, asthma and inflammatory conditions, and associated cardiometabolic markers. Future work should test these mechanisms across relevant tissues and hormonal life stages, extend analyses to more diverse populations, and evaluate whether integrating sex-specific biological and behavioural profiles can improve prevention and clinical management of cardio-psychiatric comorbidity.

## Acknowledgements

C.R., J.S., D.P. and S.v.d.S were funded by NWO Gravitation: BRAINSCAPES: A Roadmap from Neurogenetics to Neurobiology (grant no. 024.004.012 [to D.P.]). A.A.S., N.P., S.E.S., L.R., D.vd.M., K.S.O’C., and O.A.A. were funded by the Research Council of Norway (273291□[to O.A.A.], 273446□[to O.A.A.], 296030□[to O.A.A.], 324252□[to O.A.A.], 324499□[to O.A.A.], 326813□[to O.A.A.]) and European Union’s Horizon 2020 Research and Innovation Programme (Grants 847776 [CoMorMent; to O.A.A.] and 964874 [RealMent; to O.A.A.]). S.E.S. is funded by the Novo Nordisk Foundation (NNF23OC0099658 [to S.E.S.]). D.P. was funded by a European Research Council advanced grant (no. ERC-2018-AdG GWAS2FUNC 834057 [to D.P.]). EvW is financially supported by funds of the Academic Education and Research sector plans of the Dutch Ministry of Education, Culture and Science.

This work was partly performed on the TSD (Tjeneste for Sensitive Data) facilities, owned by the University of Oslo, operated and developed by the TSD service group at the University of Oslo, IT-Department (USIT). Computations were also carried out on the Genetic Cluster Computer, which is financed by the Netherlands Scientific Organization (NWO: 480-05-003), by the VU University, Amsterdam, the Netherlands, and by the Dutch Brain Foundation, and is hosted by the Dutch National Computing and Networking Services SurfSARA. This research has been conducted using the UK Biobank resource under application number 16406. We thank the numerous participants, researchers, and staff from many studies who collected and contributed to the data. We particularly express our gratitude to all UKB, All of Us, iPSYCH, and PGC participants that have been so generous to share their data for analysis. Graphical abstract was Created in BioRender https://BioRender.com/mg9ycb5

## Author Contributions

C.R.: Conceptualisation, Methodology, Formal Analysis, Writing, Visualisation, Project Administration. A.A.S.: Formal Analysis, Methodology, Feedback. N.P.: Supervision, Feedback. S.E.S.: Supervision, Feedback. L.R.: Supervision, Feedback. D.vd.M.: Resources, Feedback. K.S.O’C.: Resources, Feedback. E.v.W.: Feedback. J.E.S.: Resources, Feedback.

D.P.: Feedback, Methodology, Funding Acquisition, Resources. O.A.A.: Supervision, Feedback, Funding Acquisition, Resources. S.vd.S.: Supervision, Feedback, Methodology, Writing.

## Data availability statement

Sex stratified summary statistics of IHD and DEP will upon publication become available at https://ctg.cncr.nl/software/summary_statistics.

All GWAS summary statistics are based on Human Genome Build 37 (GRCh37/hg19). Precomputed LD scores and HapMap 3 reference file were obtained from: <u>Link</u>

1000 Genomes Phase 3 Project LD reference data: <u>Link</u>

Gene information was obtained from the GeneCard database (v5.12.0 Build 702; https://www.genecards.org).

SNP information was obtained from Open Target Genetics: https://genetics.opentargets.org

## Software Availability

FLAMES(v1.1.0) https://github.com/Marijn-Schipper/FLAMES

LAVA(v0.1.0) local genetic correlations: https://ctg.cncr.nl/software/lava

PLINK (v1.9) https://www.cog-genomics.org/plink/

---

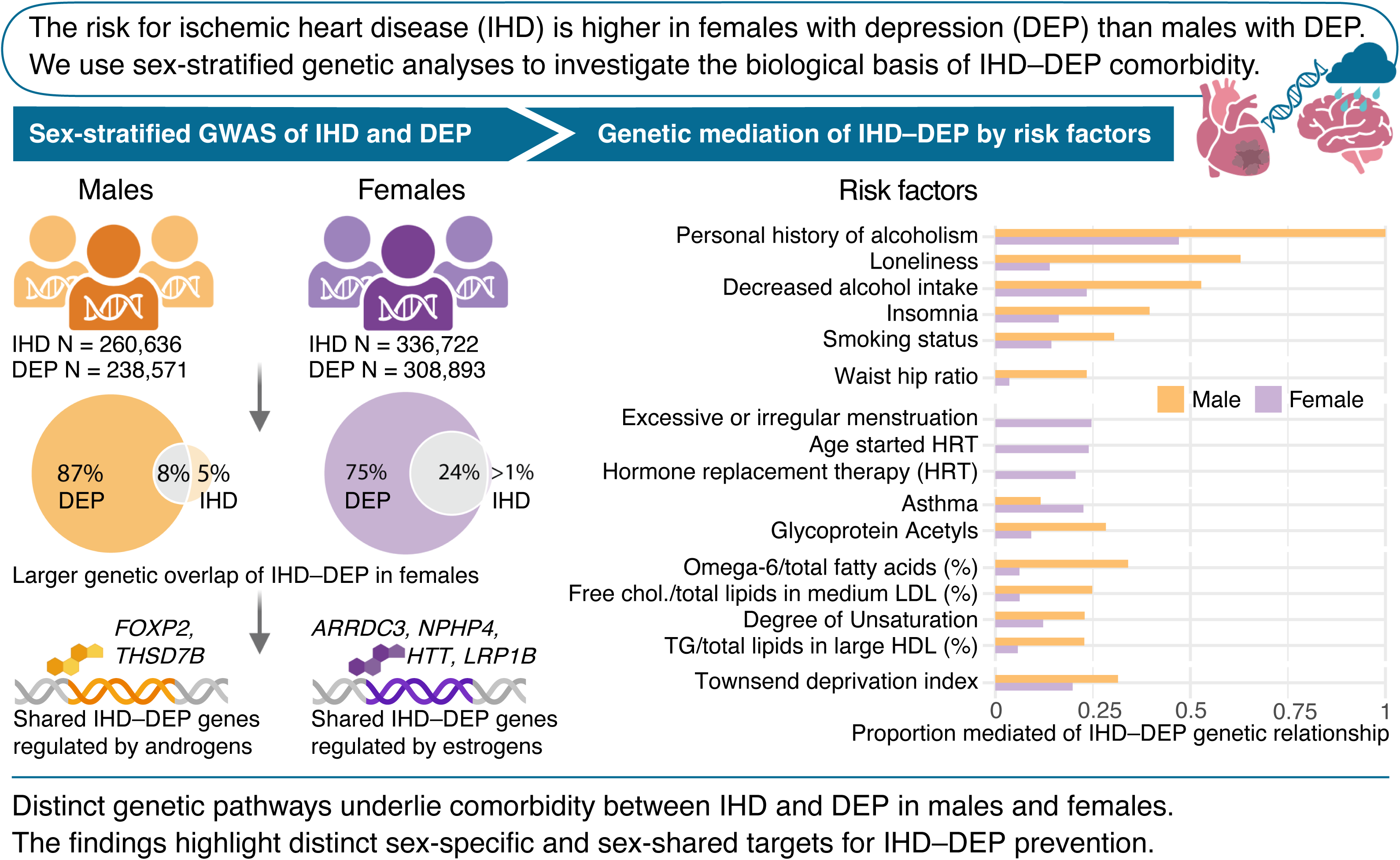

