## Supplementary Notes and Figures for "Sex differences in genetic pathways underlying ischaemic heart disease–depression comorbidity"

### **Supplementary Note 1. Cohort descriptions**

*IHD UKB sample*

Within UK Biobank, IHD cases were defined by ICD-10 codes (I21, I22, I23, I25; or equivalent ICD-9 codes) disease occurrences from UK Biobank primary care, hospital inpatient, death register, and self-reported data records (UKB data fields 1712, 3000, 2000, 40001, 40002, 20002) from the initial or follow-up visits visits to the UK Biobank assessment centers. IHD controls were defined as those without IHD diagnosis conditions . Previous EUR sex-stratified GWAS summary statistics using UKB was not available, hence we conducted the GWAS using individual level data.

*IHD All of Us sample*

Within All of Us, IHD cases were defined by systematized nomenclature of medicine clinical terms (SNOMED CT) ID 414545008, which defines IHD using a broad definition (including angina, equivalent to ICD-10 codes I20–I25) from electronic health records. IHD controls were defined as those without SNOMED ID 414545008. Previous EUR sex-stratified GWAS summary statistics using All of Us has not been conducted, hence we performed GWAS using individual level data.

*DEP UKB sample*

Sex-stratified GWAS summary statistics of DEP were obtained from Silveira et al. DEP cases were defined as individuals with either ICD-10 codes (F32, F33, F34, F38, F39) recorded across primary care, hospital inpatient, and death register (fields 1712, 3000, 2000, 40001, 40002, 20002), or by self-reported visits to a general practitioner or psychiatrist for depression (fields 2090, 2010). For additional information see Silveira et al.(1)

*DEP All of Us sample*

Within All of Us, DEP cases were defined by SNOMED ID 370143000 (equivalent to ICD-10 codes F32 and F33) from electronic health records. DEP controls were defined as those without SNOMED ID 370143000 in their electronic health records. Previous sex-stratified GWAS summary statistics using All of Us has not been conducted, hence we performed GWAS using individual level data.

*DEP PGC and iPSYCH sample*

Sex-stratified GWAS summary statistics of DEP were obtained from Blokland et al. (2022). Primary analyses combined cohorts from the Psychiatric Genomics Consortium (PGC) and the iPSYCH study in Denmark. The iPSYCH cohort included 16,438 patients with DEP and 13,538 controls, with diagnoses based on ICD-10 criteria from national register data, while PGC cohorts primarily used DSM-IV criteria. Further details on contributing cohorts, quality control, and analytic procedures are provided in Blokland et al.(2).

### **Supplementary Note 2: Sensitivity analyses for the sex difference in global genetic correlation**

*Sample overlap and phenotype definition*

Cases and controls in contributing cohorts were defined as described in Supplementary Note 1. In UKB and All of Us, controls were not explicitly excluded for IHD or DEP, meaning some degree of shared-control overlap exists across traits within the same cohort. While LDSC accounts for sample overlap via the cross-trait intercept, we assessed the possibility of residual bias of sample overlap contributing to the sex-difference in *r*_g_ by estimating the IHD–DEP *r*_g_ between the IHD GWAS meta-analysis and the independent PGC DEP cohort. The PGC DEP GWAS is clinically ascertained for DEP and does not contain UKB participants and is based on DSM-IV diagnoses. This analysis simultaneously addresses two potential sources of bias: shared-sample overlap and DEP phenotype definition, as the PGC DEP phenotype reflects clinical ascertainment rather than the broader, partly self-reported definition used in UKB.

The sex difference in IHD–DEP *r*_g_ persisted in this independent analysis (female: *r*_g_ = .331, SE = .076, *p* = 1.3×10⁻⁵; male: *r*_g_ = .167, SE = .075, *p* = .027; difference = .164, SE = .107, *p* = .125; **Supplementary Note Table 1**). Although the Wald test did not reach statistical significance, this most likely reflects the reduced power of the PGC-only DEP GWAS relative to the full meta-analysis used in the primary analyses. The direction and magnitude of the effect were fully consistent with the primary results, with the female *r*_g_ approximately twice the male estimate. Because UKB and PGC are entirely independent cohorts, these results cannot be driven by shared-sample overlap within UKB, and the use of a clinically ascertained DEP phenotype indicates that the sex difference is not an artefact of the broader DEP phenotype definition.

As an orthogonal test, we estimated cross-sex *r*_g_ estimates—that is, the genetic correlation between the IHD GWAS in one sex and the DEP GWAS in the other—which are by definition computed from non-overlapping samples and therefore eliminate shared-control bias by design. Under a sample overlap- or size-driven artefact, same-sex estimates, particularly the male–male estimate in which IHD and DEP share the same UKB male control pool, would be expected to be at least as large as cross-sex estimates. Instead, the male–male *r*_g_ (*r*_g_ = .21, SE = .04) is the lowest estimate in the set, falling below both the male IHD–female DEP (*r*_g_ = .35, SE = .04) and female IHD–male DEP (*r*_g_ = .26, SE = .03) cross-sex estimates, which are themselves closer in magnitude to the female–female estimate (*r*_g_ = .43, SE = .04). This pattern is inconsistent with inflation due to shared controls or sample overlap and instead reflects that the genetic effects underlying IHD–DEP comorbidity are genuinely stronger in females.

#### *Case-control imbalance and effective sample size*

There is a marked sex difference in the IHD-to-DEP case ratio: approximately 1:1 in males and 1:3 in females. To assess whether this imbalance in effective sample size or case-control composition could contribute to the observed sex difference in *r*_g_, we ran a sex-stratified IHD GWAS within UKB in which female and male case and control sample sizes were equalised by downsampling the IHD GWAS to match in number of cases and controls between male and female GWAS.

The sex difference in IHD–DEP *r*_g_ persisted in the downsampled analysis (female: *r*_g_ = .399, SE = .060; male: *r*_g_ = .220, SE = .059; difference = .179, SE = .084, *p* = .033; **Supplementary Table 1**), with the Wald test reaching nominal significance. These results indicate that differences in effective sample size and case-control composition do not explain the observed sex difference.

#### *IHD phenotype heterogeneity*

To evaluate whether the sex difference in *r*_g_ depends on the IHD phenotype definition, we estimated *r*_g_ between sex-stratified IHD subtype GWASs and the full DEP meta-analysis. Subtypes were defined in UKB based on individual ICD-10 codes that had sufficient statistical power for LDSC analysis, resulting in acute myocardial infarction (MI; UKB; ICD-10 I21) and chronic IHD (cIHD; UKB; ICD-10 I25) being tested. Summary statistics were obtained from the Neale lab UKB release 2.

The sex difference in *r*_g_ was directionally consistent across all IHD subtypes (**Supplementary Table 1**). For cIHD, the difference reached nominal significance (female: *r*_g_ = .328, SE = .069; male: *r*_g_ = .151, SE = .046; difference = .177, SE = .083, *p* = .033). For acute MI, the same directional pattern was observed (female: *r*_g_ = .270, SE = .088; male: *r*_g_ = .116, SE = .066), though the Wald test did not reach significance (*p* = .159), likely reflecting the reduced power of the acute MI subtype GWAS. Across all IHD subtypes, the sex difference is consistent in direction and broadly comparable in magnitude to the primary analysis, providing no evidence that the sex difference is confined to or driven by a specific IHD phenotype definition among ICD-10 diagnoses.

#### *DEP phenotype heterogeneity and depression subtypes*

To assess whether phenotypic heterogeneity within DEP contributes to the observed sex difference, we estimated *r*_g_ between the IHD meta-analysis and sex-stratified GWASs of two depression subtypes defined by neurovegetative symptom profile: atypical depression (hypersomnia and weight gain) and typical depression (insomnia and weight loss). These subtypes index distinct biological profiles that may differ in their aetiological overlap with cardiovascular disease.

The sex difference in *r*_g_ was markedly larger for atypical than for typical depression (**Supplementary Table 1**). For atypical DEP, the female *r*_g_ with IHD was .384 (SE = .095) and the male *r*_g_ was .126 (SE = .097), yielding a difference of .257 (SE = .136, *p* = .059). For typical DEP, the sex difference was substantially attenuated (female: *r*_g_ = .220, SE = .066; male: *r*_g_ = .187, SE = .085; difference = .033, SE = .108, *p* = .761). These results indicate that the sex difference in IHD–DEP *r*_g_ is not uniform across depression subtypes and is substantially more pronounced for atypical depression. Importantly, this cannot be explained by a higher prevalence of atypical presentations among female participants alone: the *r*_g_ between IHD and atypical DEP is stronger in females than in males within the atypical subtype itself, indicating a genuinely stronger genetic overlap between IHD and atypical depression in females. This subtype specificity may reflect biological pathways of particular relevance to IHD risk in women, including those involving metabolic dysregulation, hyperphagia, and neuroendocrine function, which are more closely aligned with the atypical neurovegetative symptom profile. Larger sex-stratified GWAS of depression subtypes will be needed to clarify the extent and biological basis of this subtype-specific sex difference with adequate statistical power.

**Supplementary Note Table 1. Sensitivity analyses for the sex difference in global IHD–DEP genetic correlation**

| **IHD phenotype** | **DEP phenotype** | **Female** | | | **Male** | | | **Sex difference (Wald test)** | | |
| --- | --- | --- | --- | --- | --- | --- | --- | --- | --- | --- |
|  |  | ***r*_g_** | **SE** | **p** | ***r*_g_** | **SE** | **p** | **Δ *r*_g_** | **SE** | **p** |
| ***Sample overlap and DEP phenotype definition: IHD meta-analysis vs. clinically ascertained MDD (PGC only; Wray et al.)*** | | | | | | | | | |  |
| IHD meta-analysis | MDD (PGC) | .331 | .076 | 1.3×10⁻⁵ | .167 | .075 | .027 | .164 | .107 | .125 |
| ***Case-control imbalance and effective sample size: downsampled female IHD vs. DEP meta-analysis*** | | | | | | | | | |  |
| IHD (equal N) | MDD meta-analysis | .399 | .060 | 3.0×10⁻¹¹ | .220 | .059 | 1.8×10⁻⁴ | .179 | .084 | **.033*** |
| ***IHD phenotype heterogeneity: IHD subtypes vs. DEP meta-analysis*** | | | | | | | | | |  |
| Acute MI (UKB) | MDD meta-analysis | .270 | .088 | .002 | .116 | .066 | .079 | .154 | .110 | .159 |
| cIHD (UKB) | MDD meta-analysis | .328 | .069 | 2.0×10⁻⁶ | .151 | .046 | .001 | .177 | .083 | **.033*** |
| ***DEP phenotype heterogeneity: DEP subtypes vs. IHD meta-analysis*** | | | | | | | | | |  |
| IHD meta-analysis | Atypical DEP | .384 | .095 | 5.9×10⁻⁵ | .126 | .097 | .192 | .257 | .136 | .059 |
| IHD meta-analysis | Typical DEP | .220 | .066 | 8.5×10⁻⁴ | .187 | .085 | .028 | .033 | .108 | .761 |
| ***Cross-sex genetic correlations: evidence against sample overlap artefact (within IHD and DEP meta-analyses)*** | | | | | | | | | |  |
| IHD female | DEP female | .43 | .04 | 4.1×10⁻²⁸ | *N/A (cross-sex; non-overlapping samples)* | | | | |  |
| IHD male | DEP male | .21 | .04 | 3.2×10⁻⁰⁹ | *N/A (cross-sex; non-overlapping samples)* | | | | |  |
| IHD male | DEP female | .35 | .04 | 4.0×10⁻¹⁵ | *N/A (cross-sex; non-overlapping samples)* | | | | |  |
| IHD female | DEP male | .26 | .03 | 2.3×10⁻¹⁷ | *N/A (cross-sex; non-overlapping samples)* | | | | |  |

***Note.*** All genetic correlations (*r*_g_) were estimated using multivariate LD score regression (LDSC) in Genomic SEM; sex differences were tested using a model-based Wald test in Genomic SEM. METAL IHD = inverse-variance weighted meta-analysis of IHD GWAS from UKB and All of Us. METAL MDD = inverse-variance weighted meta-analysis of depression GWAS from UKB, All of Us, iPSYCH, and PGC. MDD (PGC) = PGC-only clinically ascertained MDD GWAS, independent of UKB. cIHD = chronic ischaemic heart disease (UKB). IHD (equal N) = female UKB IHD GWAS downsampled to match male case-control effective sample sizes. Atypical DEP = depression subtype characterised by hypersomnia and weight gain. Typical DEP = depression subtype characterised by insomnia and weight loss. Cross-sex rg estimates (bottom section) are genetic correlations between sex-stratified IHD and DEP GWASs from non-overlapping samples; a Wald test sex difference comparison is not applicable (N/A) for these analyses. SE = standard error. * p < .05 (Wald test). For lower-powered analyses, non-significant Wald test p-values should be interpreted in light of the direction and magnitude of the point estimate difference rather than as evidence of equivalence.

### **Supplementary Note 3. Detailed example and explanation of conditional analysis**

The goal of the conditional genetic analysis is to assess whether the genetic overlap between two traits involves the genetic signal of a third trait. This design is adapted from mediation analysis. Mediation analysis investigates whether the correlation between two traits is induced by their joint association with a third, independent trait, i.e., a mediator.

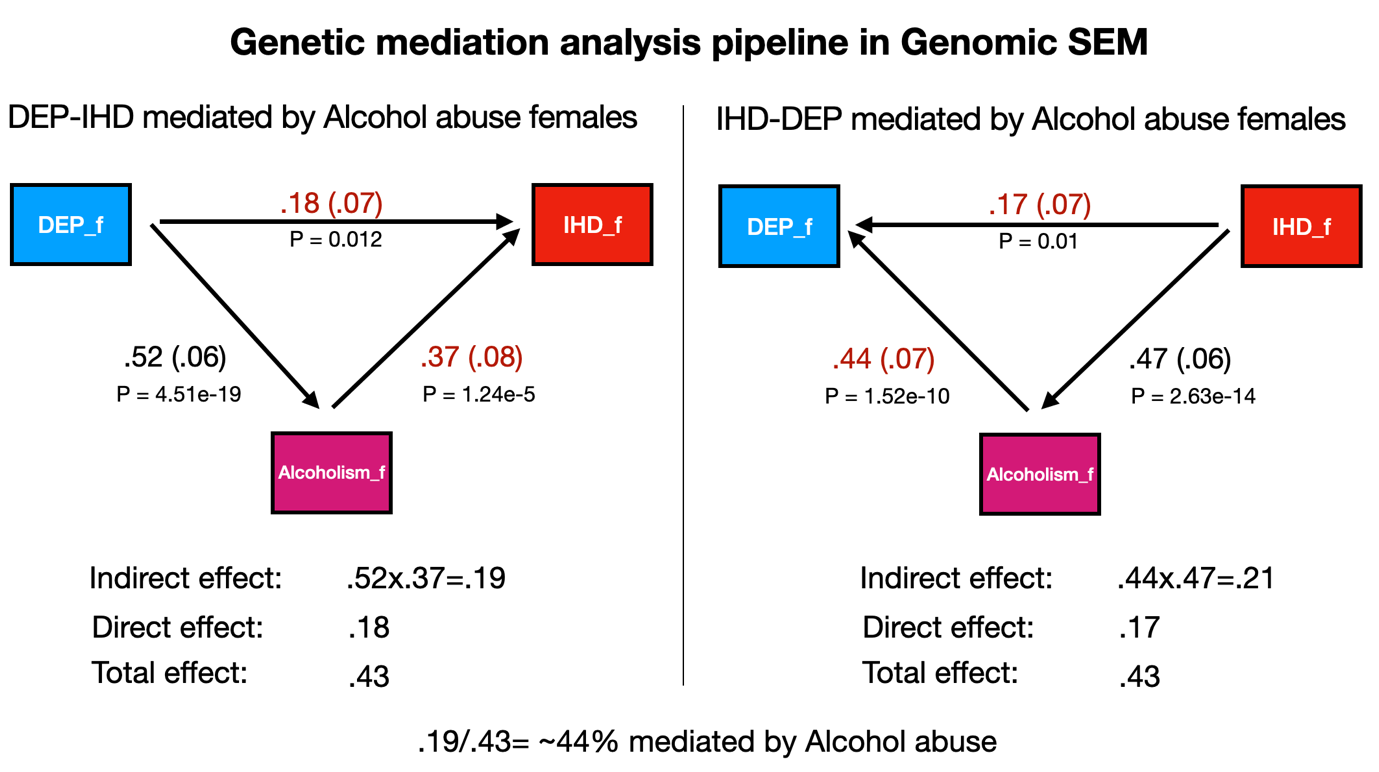

**Supplementary Note Figure 1. Illustration using alcohol abuse in females**

As an example, we tested alcohol abuse as a covariate in the genetic correlation (*r*_g_) between IHD and DEP in females (**Supplementary Note Figure 1**). The uncorrected (marginal) association between IHD and DEP was .43. After correcting for alcohol abuse, the association dropped to .17–.18, depending on whether IHD or DEP was set as the outcome. The association after controlling for alcohol abuse is the direct effect. Including alcohol abuse revealed that part of the IHD-DEP overlap ‘runs through’ alcohol abuse. This is the indirect effect, calculated as the product of two associations: alcohol abuse with DEP (.52) and alcohol abuse with IHD (.37). Their product is .19. To interpret this, we divided .19 by the total effect (.43). This shows that 44% of the IHD-DEP association is mediated by alcohol abuse.

The significance of this reduction can be tested in two steps. First, predictors (e.g., alcohol abuse and DEP) must be significantly correlated. Second, the *P*-value of the association between the covariate and the outcome (while conditioning on the other predictor) indicates whether the reduction is significant. For a derivation of this, see Romero et al. (3). Because the indirect effect is a product of two independent associations, it has no straightforward standard error. Twisk (2024; (4)) suggests using the standard error of the direct effect for visualization.

Sometimes the choice of outcome (i.e., IHD→DEP, DEP→IHD) strongly affects the mediation estimate. Mediation tends to appear larger when the outcome is the trait most strongly correlated with the covariate, and smaller when the model is reversed. Such results may reflect the strong covariate-trait link rather than true mediation. To avoid this bias, we estimated models with both IHD and DEP as outcomes and reported the result with the smaller indirect effect. This conservative approach focuses on more robust mediation signals.

### **Supplementary Note 4. Detailed discussion of prioritized genes from male and female results**

Among the female-prioritized genes, several lines of evidence indicate hormone-responsive regulation with potential relevance for IHD–DEP comorbidity.

*LRP1B* (Low-density lipoprotein receptor-related protein 1B) expression is primarily found in the brain (with differentially spliced form present in the adrenal gland and in the testis)(5) and has been shown to differ between males and females within specific inhibitory neuron subtypes regulated by estrogen-linked chromatin accessibility(6). *LRP1B* has a neuroprotective effect by reducing amyloid beta toxicity through *APP*(7,8), and reduction in estrogen following menopause could unmask this effect by lowering *LRP1B* expression(9). Knock-out mouse models without *LRP1B* display increased food intake, in addition to a female-specific phenotype of increased circulating iron levels(10), which is increasingly linked to both IHD(11) and DEP(12) risk. Previous GWAS studies have implicated *LRP1B* in vaginal microbiome measurement, breastfeeding duration, age at menarche, neuroticism, alcohol consumption quality, preeclampsia and cardiovascular disease(13).

*HTT* (Huntingtin) expression in the brain is regulated by estradiol and is required for an estrogen-linked neuroprotective program involving neuroglobin trafficking to mitochondria under oxidative stress(14). Similarly to *LRP1B*, declining estrogen around menopause could diminish *HTT*-related resilience. Additionally, knock-out mice models show female-specific phenotypes such as hyperactivity, excessive grooming behavior and decreased body weight(10). *HTT* CAG repeat size variations have been linked to cortical thickness only in females(15), and to cardiac disorders and adverse lipoprotein profiles in both sexes(16,17). Previous GWAS studies have implicated *HTT* in BMI, alcohol consumption, worry, and DEP(13).

*ARRDC3* (Arrestin Domain Containing 3) has been shown to be regulated by estrogen in the liver(18) where it participates in insulin modulation and glucose metabolism. *ARRDC3* acts as an α-arrestin adaptor for β-adrenergic receptors and shows a sex-dependent impact on obesity and energy expenditure(19). During pregnancy, *ARRDC3* methylation affects growth trajectories only in female offspring(20). *ARRDC3* further participate in angiogenesis and pathology of preeclampsia(21)—a female-specific condition associated with higher later-life cardiovascular risk(22) and mood vulnerability(23).

*NPHP4* encodes a ciliary transition-zone protein that regulates Wnt/Shh signaling at the ciliary transition zone(24,25). This pathway is central to ovarian follicle development, luteinization, and steroidogenesis(26). Concordant with this, *NPHP4* has been linked to ovarian dysfunction, placenta retention and placenta abruption(27). In addition, *NPHP4* has been linked to heart malformations(28), cardiovascular events(29), IHD risk(30), and neuroticism(31). Together, these female-prioritized loci suggest that shared liability between IHD and DEP in females may be more strongly shaped by hormone-responsive, reproductive, and metabolic pathways.

By contrast, the male-prioritized loci align more closely with androgen-linked neurodevelopmental and behavioral liability. *FOXP2* (Forkhead box P2) encodes a forkhead transcription factor with roles in neural and cardiopulmonary development and synapse regulation(32,33). *FOXP2* expression in the brain shows sex differences in a region-specific manner(34) potentially due to upstream regulatory effects of androgens(35). Knock-out mouse models without *FOXP2* show increased pulmonary respiratory rate, abnormal eating behavior, abnormal cardiovascular system physiology, decreased QRS amplitude, and increased dopamine and serotonin levels(36). *FOXP2* is further implicated in sinoatrial node pacemaker cells development(37) and has been associated with DEP and schizophrenia(38). Androgen-responsive *FOXP2* programs could influence male-leaning cortical/autonomic features while linking neural and cardiac biology relevant to IHD–DEP comorbidity. GWAS studies link *FOXP2* to apraxia, ADHD, insomnia, and risk taking behavior(13).

*THSD7B* (Thrombospondin Type 1 Domain Containing 7B) participates in actin cytoskeleton organization and has been implicated in angiogenesis and prostate cancer(13). Knock-out mouse models without *THSD7B* display male-specific phenotypes including decreased body length and increased circulating triglyceride levels(10), and *THSD7B* expression is differentially regulated by androgens(39). Human genetic studies have linked *THSD7B* with smoking initiation, neuroticism, and metabolic traits such as BMI and lipid levels(13), and specific SNPs in *THSD7B* have been linked to greater ethanol-induced adverse effects(40–42).

### **Supplementary Figures 1-9.**

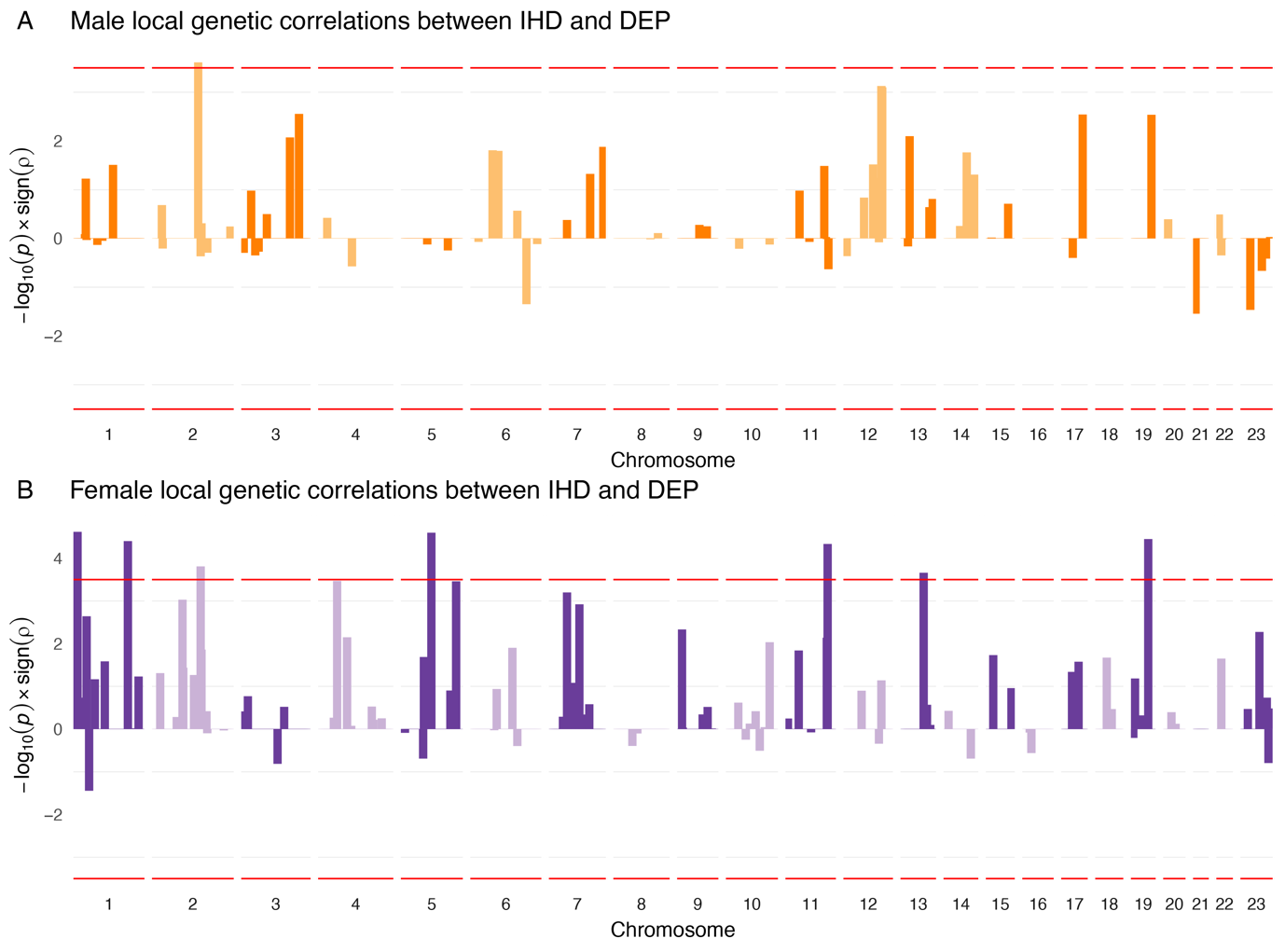
**Supplementary Figure 1. Local genetic correlations in LAVA between IHD and DEP in males and females**

*Note:* *Local r*_g_ *between IHD and DEP over all loci in the genome in A) males (orange) and B) females (purple). The y-axis is defined as −log10(P) × sign of rho. Dotted red bar represents Bonferroni significance threshold (α_BON_ = 3.16 × 10^−4^).*

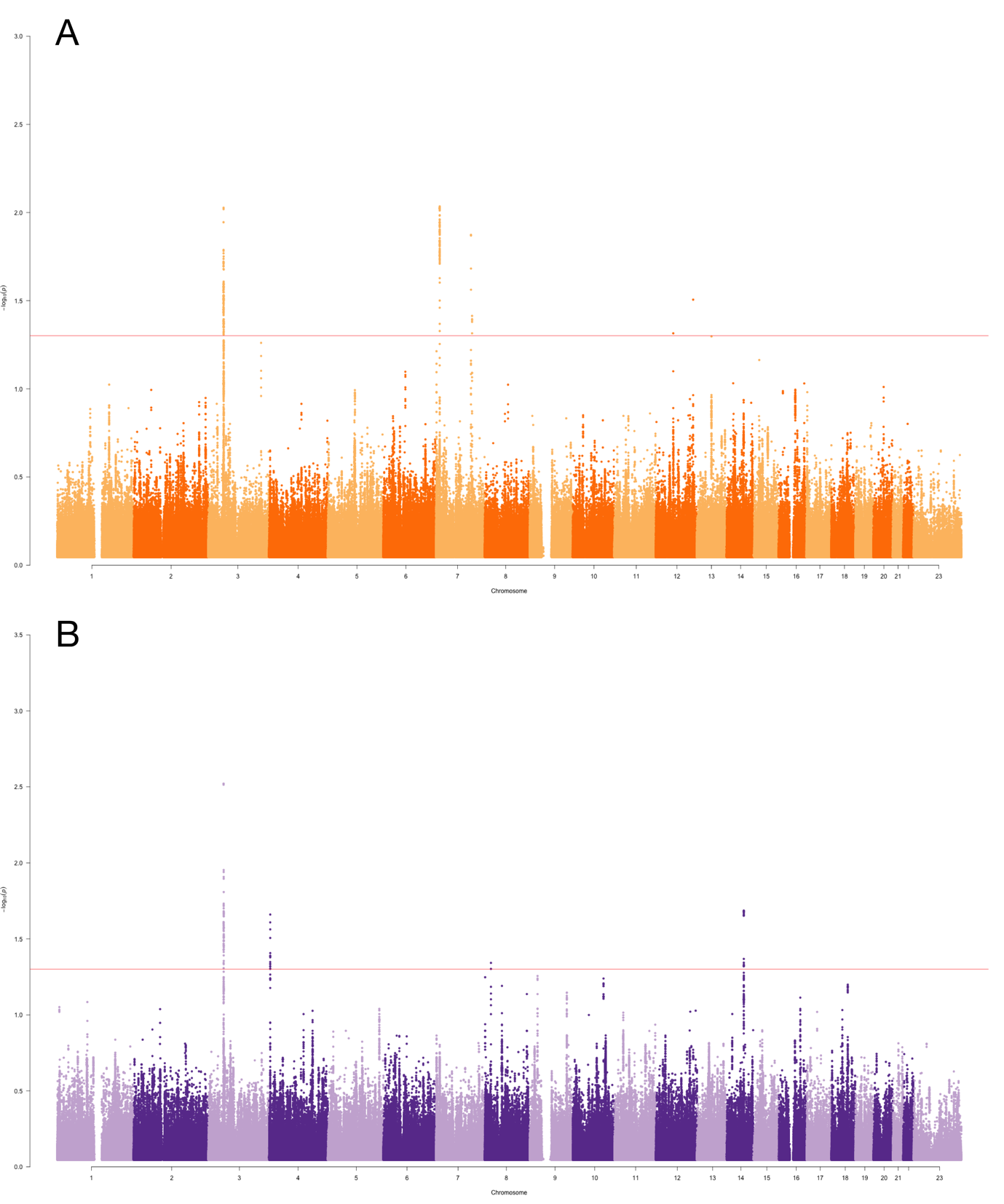
**Supplementary Figure 2. SNPs jointly associated with IHD and DEP**

*Note: The Manhattan plots show SNPs jointly associated with IHD and DEP at the conjunctional false discovery rate < 0.05 (red line) for males (A) orange) and females (B) purple). The y-axis is defined as −log10(conjFDR) for each SNP and chromosomal positions along the x-axis.*

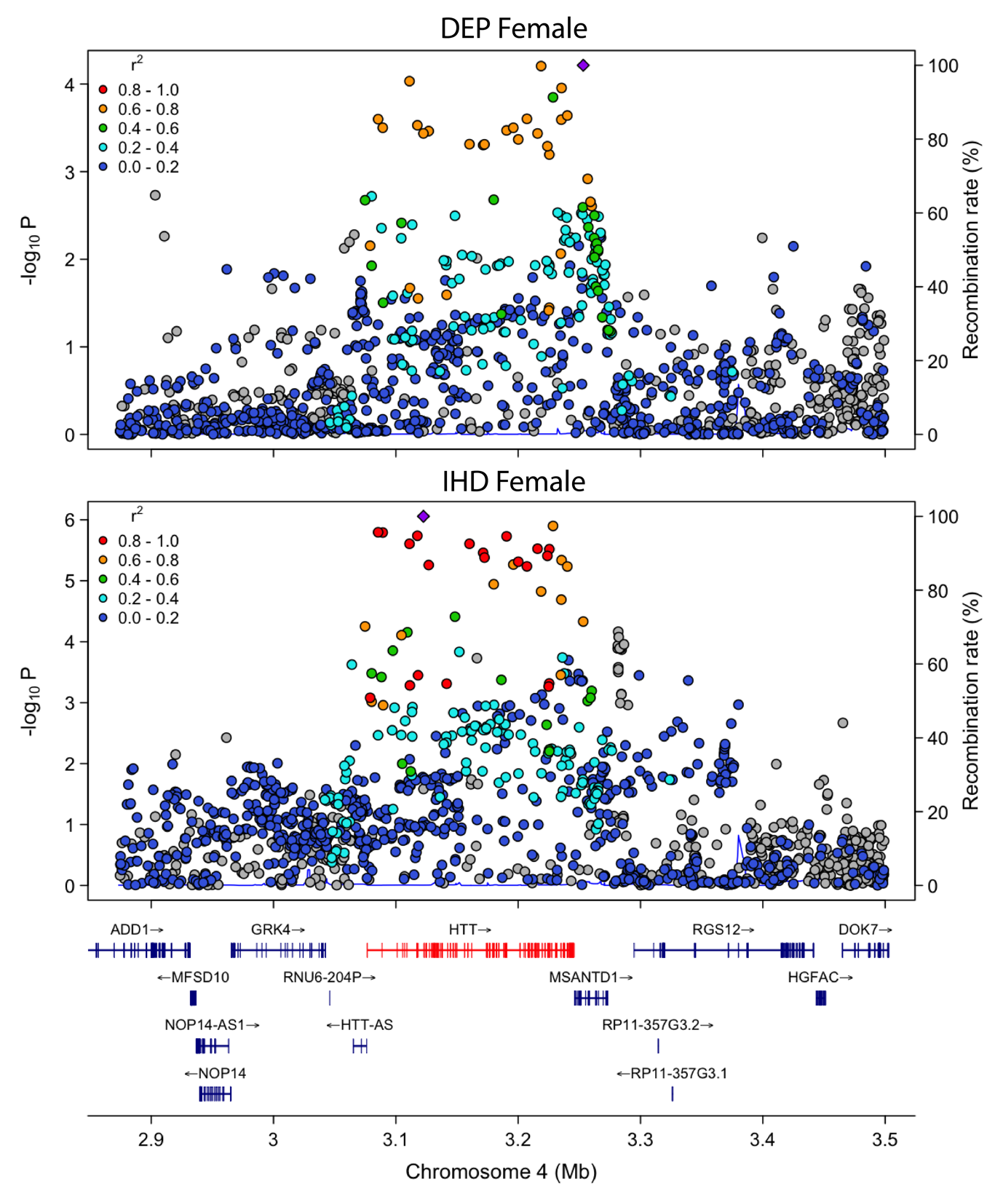

#### **Supplementary Figure 3. Locus zoom plot *HTT* genomic region in females**

*SNPs are dots in locus zoom plot and colored by their LD R^2^ value with reference to the most significant SNP in the locus. Plots are shown for the same locus for DEP (top) and IHD (bottom). Generated using LDlink.*

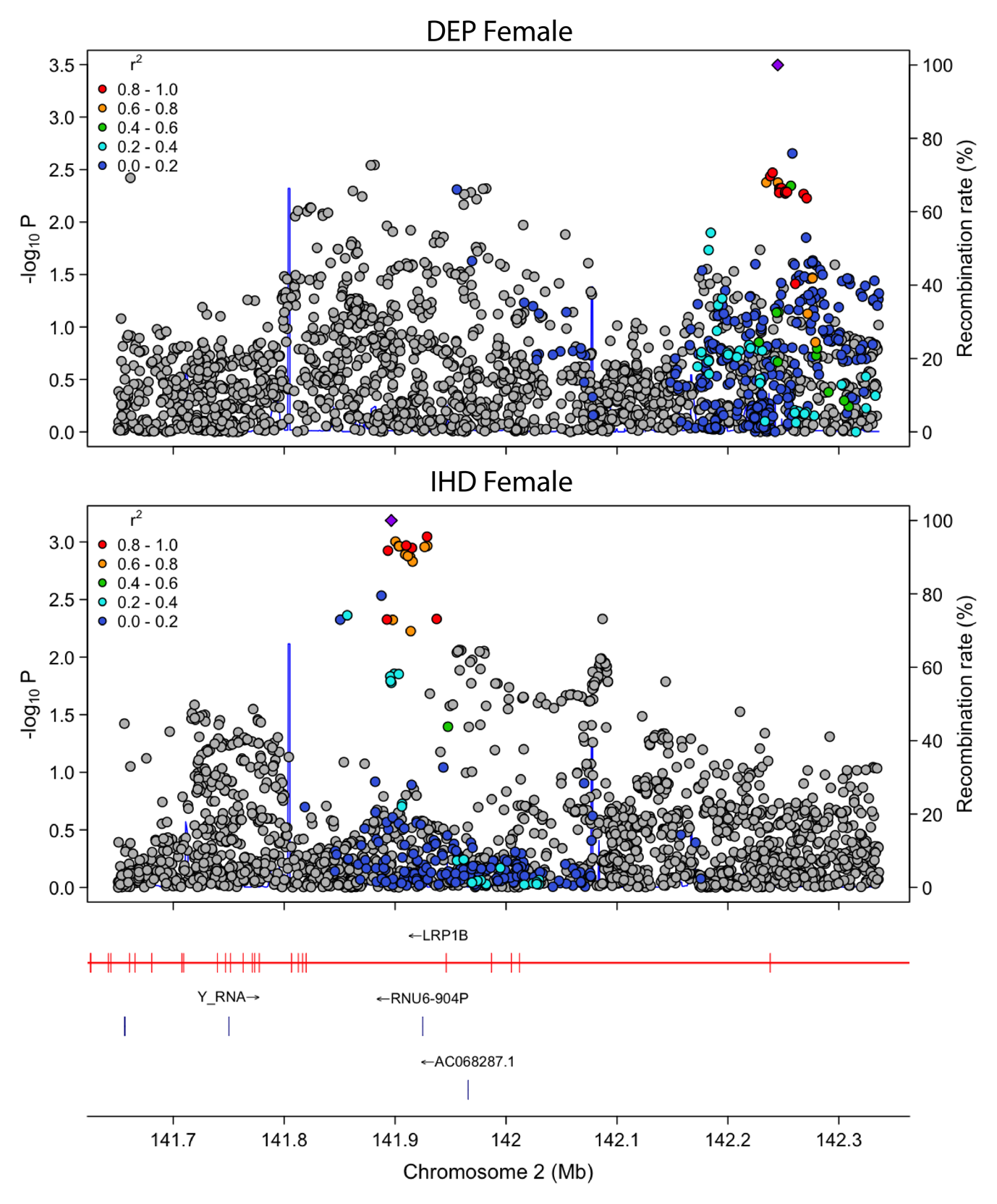

#### **Supplementary Figure 4. Locus zoom plot *LRP1B* genomic region in females**

*SNPs are dots in locus zoom plot and colored by their LD R^2^ value with reference to the most significant SNP in the locus. Plots are shown for the same locus for DEP (top) and IHD (bottom). Generated using LDlink.*

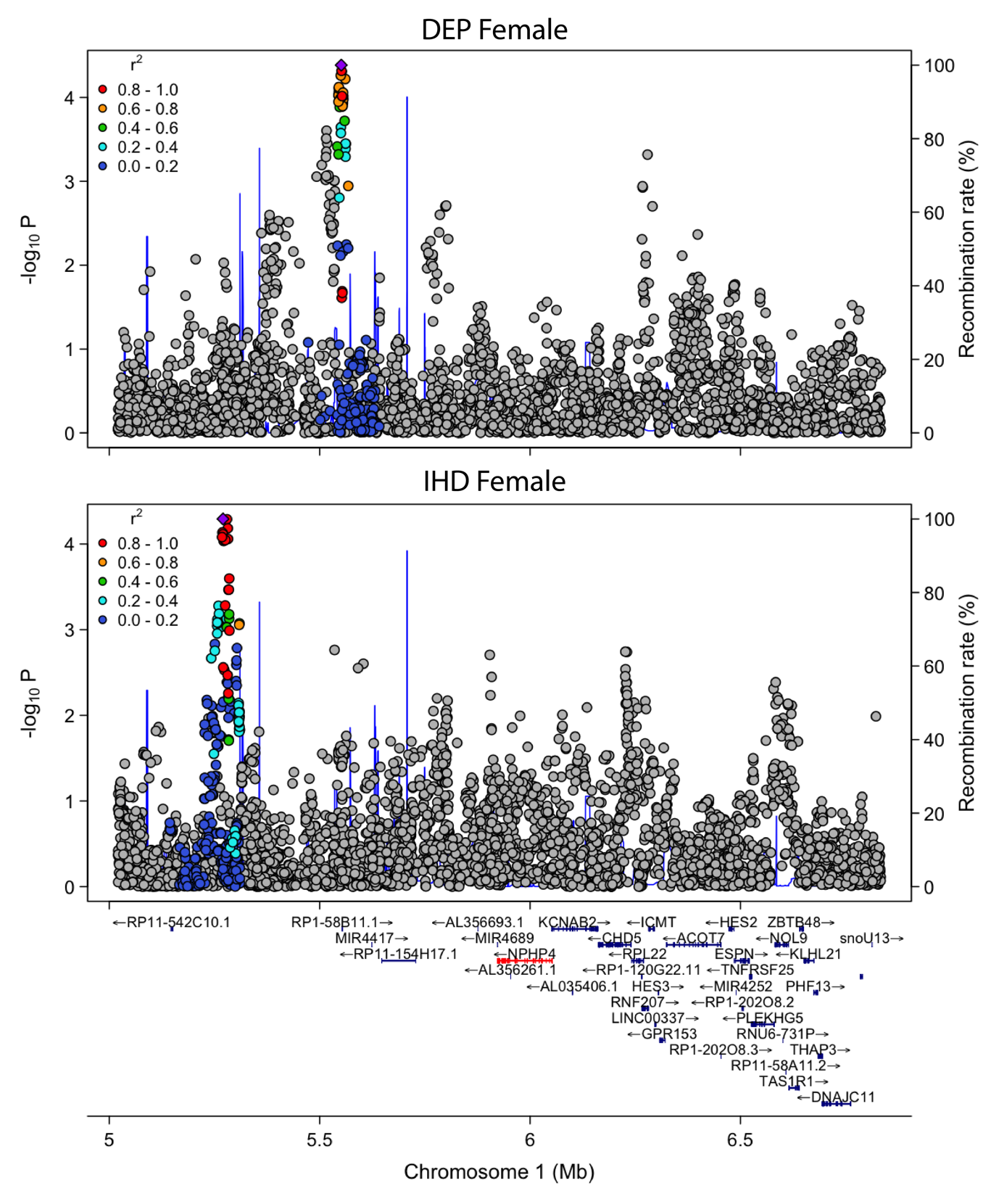

#### **Supplementary Figure 5. Locus zoom plot *NPHP4* genomic region in females**

*SNPs are dots in locus zoom plot and colored by their LD R^2^ value with reference to the most significant SNP in the locus. Plots are shown for the same locus for DEP (top) and IHD (bottom). Generated using LDlink.*

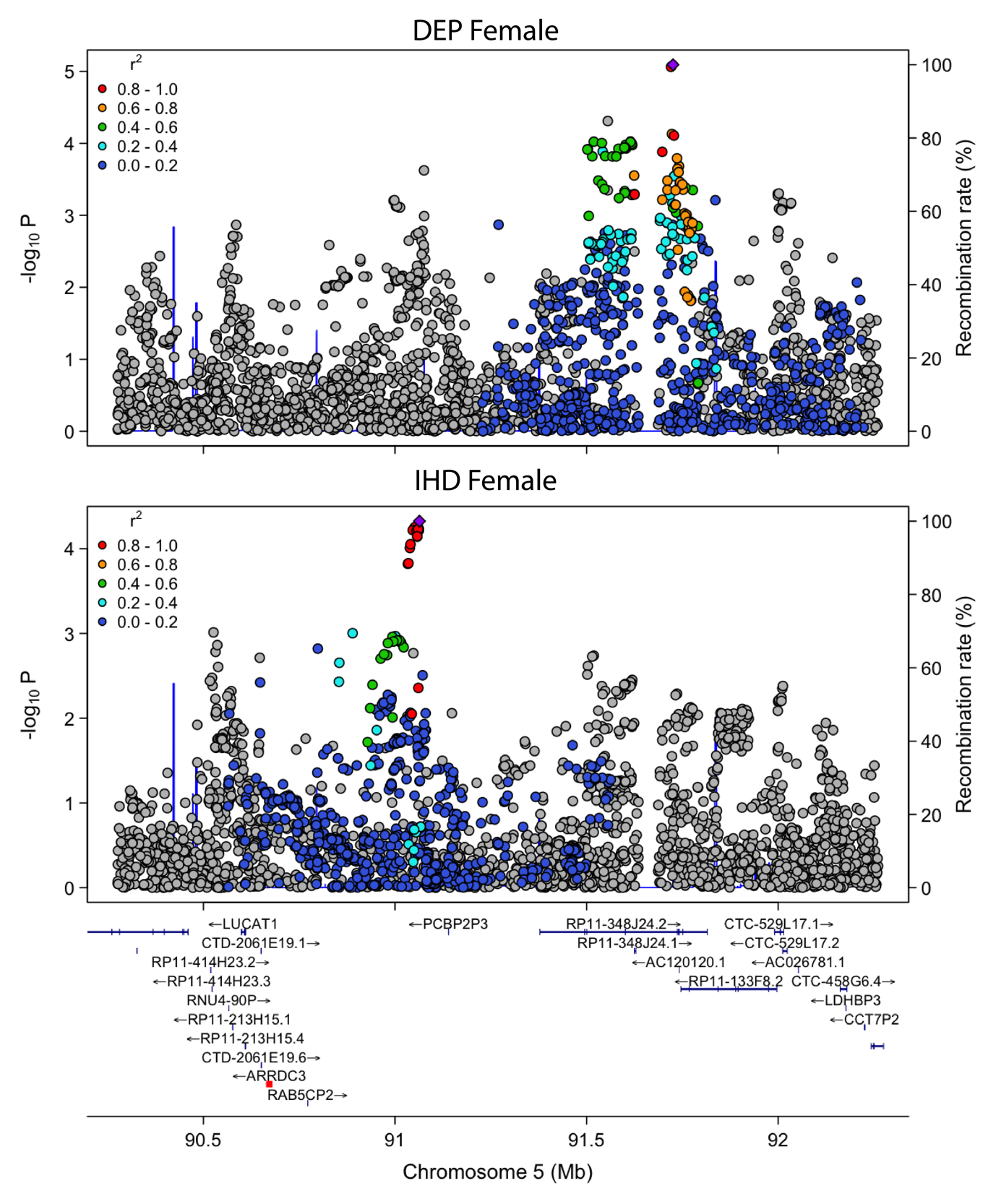

#### **Supplementary Figure 6. Locus zoom plot *ARRDC3* genomic region in females**

*SNPs are dots in locus zoom plot and colored by their LD R^2^ value with reference to the most significant SNP in the locus. Plots are shown for the same locus for DEP (top) and IHD (bottom). Generated using LDlink.*

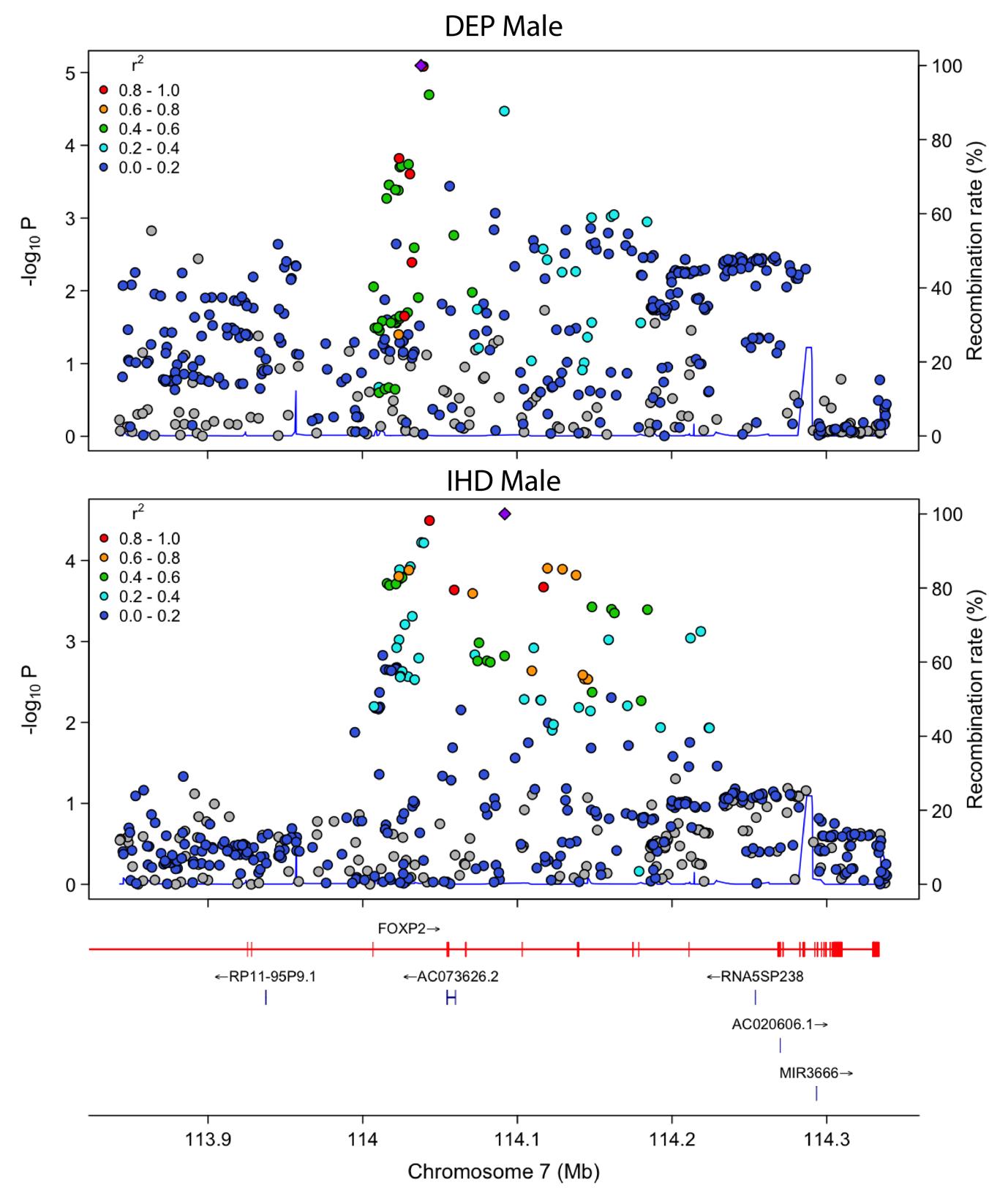

#### **Supplementary Figure 7. Locus zoom plot *FOXP2* genomic region in males**

*SNPs are dots in locus zoom plot and colored by their LD R^2^ value with reference to the most significant SNP in the locus. Plots are shown for the same locus for DEP (top) and IHD (bottom). Generated using LDlink.*

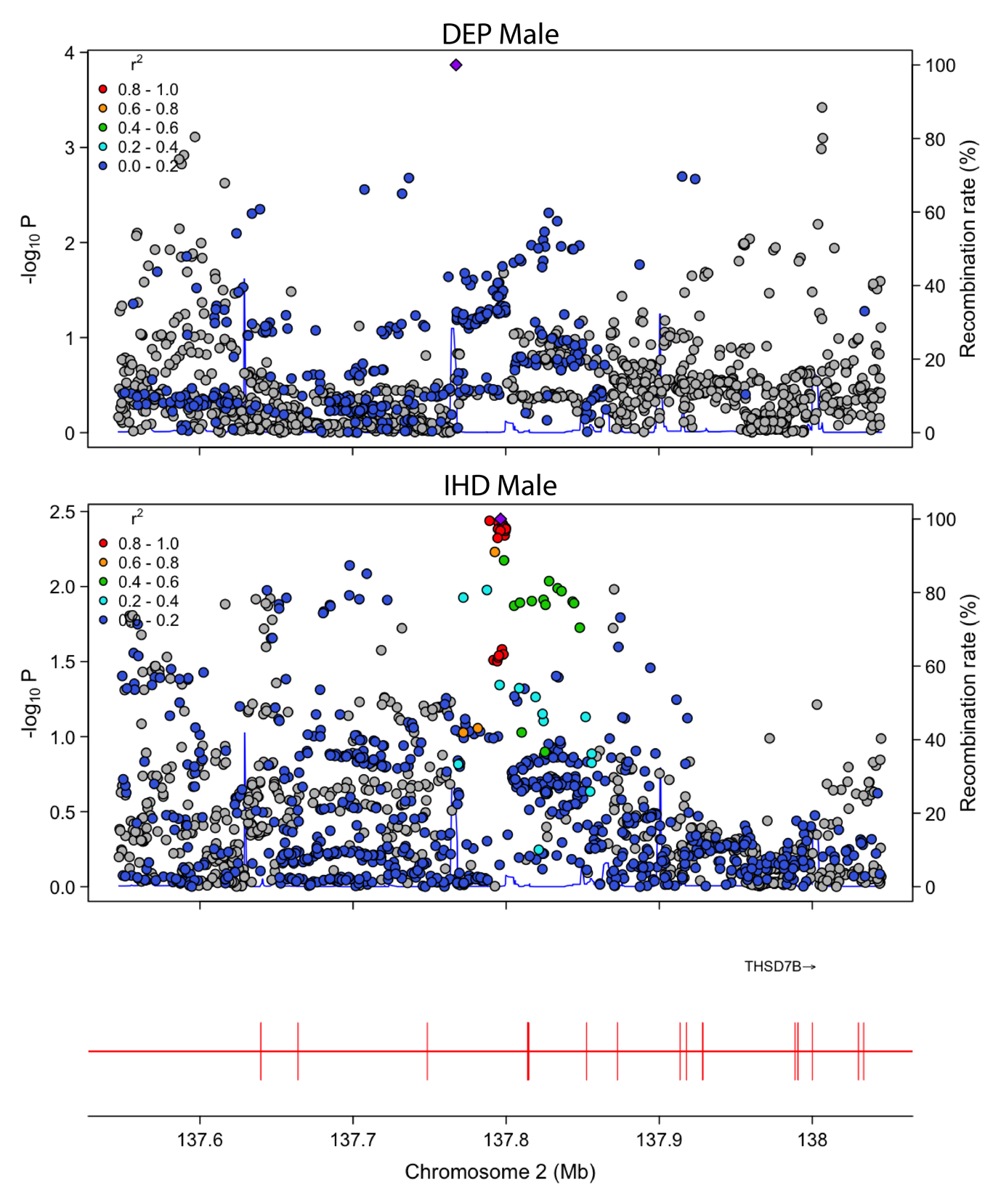

#### **Supplementary Figure 8. Locus zoom plot *THSD7B* genomic region in males**

*SNPs are dots in locus zoom plot and colored by their LD R^2^ value with reference to the most significant SNP in the locus. Plots are shown for the same locus for DEP (top) and IHD (bottom). Generated using LDlink.*

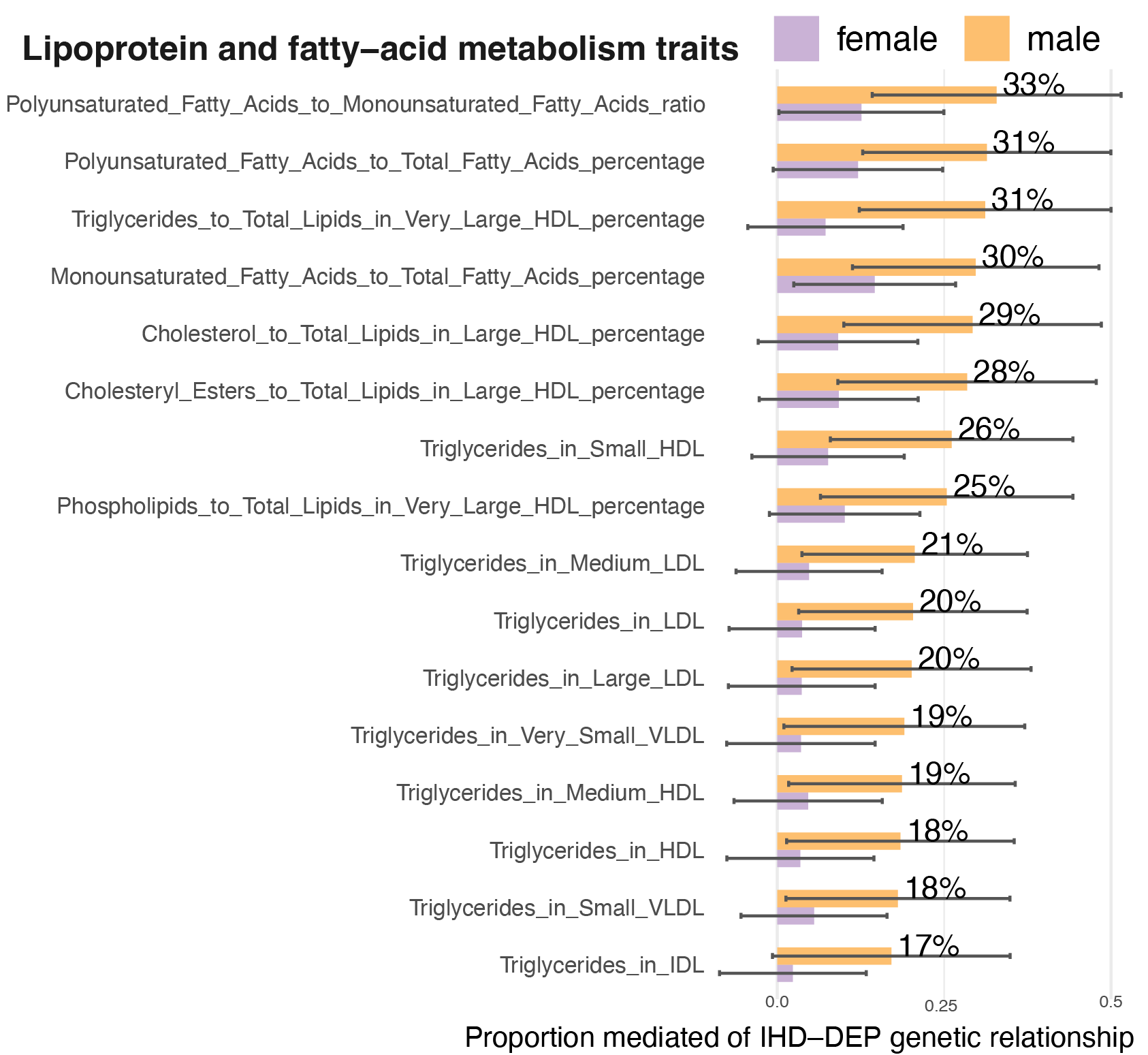

**Supplementary Figure 9. Genetic mediation results from all blood measurements**

*Significant genetic mediator traits in either sex are plotted for males (orange) and females (purple). The y-axis shows the proportion mediated, defined as the ratio of the indirect effect of the mediator to the marginal IHD-DEP r*_g_*. For each mediator, mediation was estimated in both model orientations (IHD→DEP and DEP→IHD), and the minimum proportion mediated is plotted as a conservative estimate. Standard errors of the indirect effects were approximated using the standard errors of the corresponding direct effects, as recommended (4). NS indicates non-significant mediation.*
